# Allocation of COVID-19 Vaccines Under Limited Supply

**DOI:** 10.1101/2020.08.23.20179820

**Authors:** Xin Chen, Menglong Li, David Simchi-Levi, Tiancheng Zhao

## Abstract

**Problem definition:** This paper considers how to allocate COVID-19 vaccines to different age groups when limited vaccines are available over time.

**Academic/practical relevance:** Vaccine is one of the most effective interventions to contain the ongoing COVID-19 pandemic. However, the initial supply of the COVID-19 vaccine will be limited. An urgent problem for the government is to determine who to get the first dose of the future COVID-19 vaccine.

**Methodology:** We use epidemic data from New York City to calibrate an age-structured SAPHIRE model that captures the disease dynamics within and across various age groups. The model and data allow us to derive effective static and dynamic vaccine allocation policies minimizing the number of confirmed cases or the numbers of deaths.

**Results:** The optimal static policies achieve a much smaller number of confirmed cases and deaths compared to other static benchmark policies including the pro rata policy. Dynamic allocation policies, including various versions of the myopic policy, significantly improve on static policies.

**Managerial implications:** For static policies, our numerical study shows that prioritizing the older groups is beneficial to reduce deaths while prioritizing younger groups is beneficial to avert infections. For dynamic policies, the older groups should be vaccinated at early days and then switch to younger groups. Our analysis provides insights on how to allocate vaccines to the various age groups, which is tightly connected to the decision-maker’s objective.

## 1. Introduction

The pandemic COVID-19 has caused tremendous loss to the global economy since December 2019. To contain this pandemic, over 165 vaccines are developing, among which 37 vaccines are in human trials (Corum et al. 2020). Most experts believe that a vaccine is likely to become available by mid-2021, about 12-18 months after COVID-19 first emerged (Gallagher 2020). However, the early COVID-19 vaccine supplies are far from enough for everyone, even for people at high risk such as healthcare workers (Loftus 2020). In order to have early access to COVID-19 vaccines, the US government has made an agreement with Pfizer Inc. and BioNTech SE to provide 100 million doses of COVID-19 vaccines once proved to work safely (Wack 2020). An important question is how to allocate limited vaccines to different groups of people when it becomes available over time (Shipman 2020, Sun 2020). It is commonly agreed that highest-risk medical, national security, and other essential workers should get the vaccine first (Sun 2020). A challenging problem is how to allocate vaccines for the remaining large population.

In this paper, we focus on vaccine allocation policies for general population grouped by ages. We follow the Centers for Disease Control and Prevention (CDC) to classify people into 5 age groups: 0-17, 18-44, 45-64, 65-74, 75+. The epidemiological model we use is an age-structured SAPHIRE model whose single population version is proposed by Hao et al. (2020). See Zhou et al. (2019) and the reference therein for more on age-structure in epidemic models. Our model has seven compartments: susceptible compartment, exposed compartment, presymptomatic infectious compartment, unascertained infectious compartment, ascertained infectious compartment, isolated compartment, and removed compartment. Each compartment is further divided into five age groups. We use the COVID-19 data of New York City from March 17 to June 8 to estimate parameters in our model including the contact matrix and the ascertainment rate. The parameter estimation method we use is the standard least squares method, i.e., to minimize the squared errors of the prediction made by the epidemic model. Since the disease dynamics are nonlinear, the accumulated squared error is a highly nonlinear and nonconvex function of the parameters, which leads to many different local optimal solutions with close objectives. Due to this computationally challenge, papers in the literature often use off-the-shelf estimates from other social science papers, e.g., the seminal paper Medlock and Galvani (2009) uses the survey-based contact data from Mossong et al. (2008). Instead, we directly estimate the contact matrix using the actual data from NYC which should provide a more convincing evaluation of allocation policies. To overcome the difficulty, we impose reasonable conditions on the mixing pattern of age groups, which allows us to find the the local optimum that is most likely to be the global optimum.

Based on the estimated parameters, we solve the optimal static allocation policies under different objectives and different settings of daily supply. The objectives we consider include the total number of confirmed cases, the total number of deaths, and their weighted sum. When the daily supply is constant, our numerical study shows that to minimize the total deaths, it is optimal to allocate limited vaccines to the oldest group first and then the younger group if there are capacities remaining. When the constant daily supplies increase, the older group still gets the majority of the vaccines while the younger groups gradually get more. The optimal static policy that minimizes the total confirmed cases follows a similar pattern but allocates doses to younger groups even if the daily available vaccines are limited. When the constant daily supplies increase, the younger groups get the majority of the vaccines. We compare these two optimal static polices with two benchmark policies: a uniform allocation policy that allocates available vaccines equally to each age group, and a pro rata policy that allocates available vaccines proportionally to the population of each age group. Our numerical study shows that the optimal static policies have a significant decrease in total confirmed cases and deaths compared to the two benchmarks. Under the setting that the daily supply is increasing at the beginning and then becomes steady, the allocation pattern of optimal static policies is almost the same as that under the constant daily supply setting. We would like to point out that the analysis of optimal static vaccine allocation policy under the multi-period setting is not found in the literature.

Several dynamic allocation policies are evaluated including an old-first policy which allocates daily supplies in a decreasing age order, an infection-first policy which allocates daily supplies proportionally to the current infection ratio of each group, a myopic policy which determines today’s allocation by minimizing tomorrow’s total new infections of all groups, a death-weighted myopic policy, a two-day myopic policy which determine the allocation for today and tomorrow by minimizing the total new infections of the day after tomorrow, and a seven-day myopic policy which determine the allocation for one week by minimizing the total new infections after one week. Under the setting of constant daily supply, our numerical study shows that the three types of myopic policies have similar outcomes and significantly outperforms the other dynamic allocation heuristics and optimal static policies in terms of both confirmed cases and deaths. The myopic policy allocates daily available vaccines to the group with the largest marginal vaccination effect, which leads to a single-group allocation. The two-day and seven-day myopic policies have mixing allocations over groups but still follow a similar pattern with the myopic policy in that the group that gets the majority of the daily supply is roughly the same with the group that gets all under the myopic policy. This suggests the decision-maker to take the marginal vaccination effect of each group into consideration when making allocation. When the daily supply is increasing at the beginning periods and then becomes steady, all the myopic policies still perform the best while the old-first policy performs slightly worse. However, they all have significant increases in confirmed cases and deaths compared to the setting of constant daily supply. This is because there is limited room for improvement when the supply at the beginning is scare, which illustrates the importance of vaccine supply in early stages of the epidemic.

Finally, we discuss the trade-off between efficiency and equity. To account for it, we consider a policy that allocates a portion of the daily supply according to the pro rata policy and allocates the remaining portion according to the myopic policy. Our numerical study shows that when more doses are reserved for the myopic policy, both the confirmed cases and deaths averted have a decreasing margin. Moreover, 50% reduction in the averted cases associated with pure myopic policy can be achieved by reserving only 30% of the capacity for the myopic policy, and 47% reduction in averted deaths associated with pure myopic policy requires merely 20% of the capacity to the myopic policy. We also examine the impact of fairness on the myopic policy using Gini index.

The organization of the paper is as follows. Section 2 provides a literature review of papers on vaccine allocation. In Section 3, we formally introduce the age-structured SAPHIRE model. Section 4 presents the description of the data set and details of parameter estimation. Section 5 contains comprehensive numerical experiment of allocation policies. We conclude the paper in Section 6.

## 2. Literature Review

This paper is most relevant to papers on vaccine allocation for the COVID-19 pandemic. Babus et al. (2020) consider a joint COVID-19 vaccine allocation and stay-at-home order problem over different age-occupation groups. They use a fractional probit model to calculate the probability of infection given a physical proximity and form a linear program to determine the optimal vaccine allocation and stay-at-home order for age-occupation groups with an objective of minimizing the risk of infections and the economic losses. They illustrate that vaccine allocation should emphasize age-based mortality risk more than occupation-based exposure risk. This motivates our study on the allocation of COVID-19 vaccines to different age groups. Matrajt et al. (2020) use a non-interacting age-stratified SEIR based compartmental model to study the vaccine allocation problem for COVID-19 to different age groups. They set all the parameters in the model according to the literature and solve the optimization problem with a single capacity constraint under different objectives. They find that when minimizing deaths, it is optimal to vaccinate group 75+ first if the vaccine effectiveness is low, while it is optimal to vaccinate younger groups first if the effective coverage is high. When minimizing symptomatic infections, priority was given to the younger groups. In this paper, we consider multi-period allocation and allow mixing across groups. We obtain a similar characterization of the optimal static allocation policies under the above objectives. In addition, we propose several dynamic heuristics that outperforms static policies in terms of both confirmed cases and deaths. There are some papers provide guidelines for (ethical) allocation of limited doses of future COVID-19 vaccines. See Henn (2020), Emanuel et al. (2020a), Emanuel et al. (2020b), and Liu et al. (2020b). Recently, the Advisory Committee on Immunization Practices (ACIP) has a workshop discussing COVID-19 vaccine prioritization, see Dooling (2020) for more details.

This paper is related to papers studying other interventions for COVID-19. For example, Birge et al. (2020) study the priority of lockdowns for regions in NYC using a variant of space-stratified SEIR compartmental model with off-the-shelf disease transmission parameters and commuting matrix calculated with data from SafeGraph. Housni et al. (2020) use a SIR based compartmental model to study the effect on the testing capacity after reopenning.

Papers studying vaccine allocation for other diseases like influenza are closely related to this paper even though COVID-19 has different characteristics (e.g., larger mortality rate and different morbidity rate compared with influenza). Mylius et al. (2008) develop an age-structured SEIR model validated by the 1957-1958 asian flu pandemic. They compare the effectiveness of two policies, a policy prioritizing high-complication risk and a policy prioritizing high-risk of infection, and show that which one is better depends on the time of vaccination. Medlock and Galvani (2009) develop an age-structured SEIR model with additional compartments for vaccinated individuals to determine the optimal vaccine allocation for the swine-origin H1N1 influenza outbreak. Their contact matrix shows strong mixing within the same age groups and moderately high mixing between children and people of their parents’ ages, This renders the optimal allocation to prioritize schoolchildren and adults aged 30-39 as schoolchildren are most responsible for transmission and their parents serve as bridges to the rest of the population. Medlock et al. (2009) further extend Medlock and Galvani (2009) to include two levels of risk for complications due to influenza infection and incorporate staggered delivery of vaccine doses. Lee et al. (2010) use an agent-based SEIR model to justify the effectiveness of the at-risk individuals-first policy recommended by ACIP for the 2009 H1N1 influenza pandemic. Tuite et al. (2010) develop an age-structured SEIR model in which each age group is further classified by risks, and evaluate four vaccination strategies using the 2009 H1N1 data of Ontario, Canada. Matrajt and Jr (2010) use a similar model with only two age groups (children and adults) calibrated by the 2009 H1N1 pandemic in US to compare optimal vaccination strategies started at different time during the pandemic under the settings of developing country and developed country. Yarmand et al. (2014) consider a two-phase vaccine allocation problem in which a limited doses of vaccines are allocated to different regions in the first phase and additional doses are allocated in the second phase to contain the epidemic. They propose two formulations of the problem, a two-stage stochastic linear program and a newsvendor formulation, and test their solutions for the seasonal influenza in North Carolina. Lee et al. (2015) propose a disease propagation model coupled with a vaccination queuing model which can be used to derive the optimal timing for switching from the prioritized vaccination strategy to the nonprioritized strategy during the course of the influenza pandemic. Nguyen and Carlson (2016) use a space-structured stochastic SIR model to derive the optimal vaccine allocation under different time of vaccination, interacting levels between cities, and vaccine capacities. Dalgiç et al. (2017) numerically compare the effectiveness of vaccine policies derived from agent-base models and compartmental models for influenza pandemic. Duijzer et al. (2018) study a non-interacting meta-population SIR model and provide structural characterizations of the the optimal vaccine allocation. They propose a concept of does-optimal (similar the vaccination marginal effect in this paper), and show that to minimize total infections, it is inefficient to allocate vaccines to groups under post-peak stage of the epidemic. We refer to Li et al. (2018) and Venkatramanan et al. (2019) for more recent work on influenza vaccine allocation. See Moore and Lessler (2015) for the optimal allocation of oral cholera vaccines.

The control of the above papers are all one-shot allocations (except Yarmand et al. (2014) considering a two-phase allocation). To the best of our knowledge, Teytelman and Larson (2013) and Long et al. (2018) are the only two papers considering dynamic allocation policies for epidemics. Teytelman and Larson (2013) employ a new non-interacting space-structured influenza-spread model with parameters inferred from the 2009-2010 H1N1 pandemic. They propose several dynamic heuristics including a pro rata heuristic, a prepeak heurisitc, a greedy heuristic, a critical period heuristic, and a telescope-to-the-future switching algorithm heuristic. They show that the switching algorithm heuristic performs the best in terms of infections averted. It is worth pointing out that although the greedy heuristic allocates the vaccine doses one by one to the region with the highest marginal benefit for the next vaccine while the myopic policy in this paper allocates the total daily available doses to the age group with the highest vaccination marginal benefit. Moreover, since the epidemic model in this paper is totally different from the one in Teytelman and Larson (2013), the vaccination marginal benefit is computed differently. Moreover, they only provide numerical values of vaccination marginal benefits under different parameter settings while we provide an analytic form with a direct explanation.

Long et al. (2018) develop a space-structured SIR model and estimate the contact matrix by a gravity model. Using the 2014 Ebola case data from Guinea, Liberia, and Sierra Leone, they evaluate several heuristic policies for hospital beds allocation: a static allocation determined by the fraction of the accumulated infections of each region at some time, a static greedy *R*_0_ policy, a myopic LP policy, and a policy obtained from approximate dynamic programming. They particularly focus on their performances under different availability of data for parameter estimation and find that overall the myopic policy performs the best. We would like to mention that the LP formulation for their myopic policy is different from the one in this paper as we focus on vaccine allocation, and more importantly, we are able to explain the allocation pattern of the myopic policy using the vaccination marginal effect. In addition, the gravity model used in Long et al. (2018) to estimate the contact matrix is demonstrated unrealistic for COVID-19 by Li et al. (2020). Hence, as this paper aims to provide valuable suggestions on COVID-19 vaccine allocation among age groups, we directly estimate the contact matrix using the epidemic model and the data from NYC (which is challenging due to the many local optimal solutions). Moreover, we propose and evaluate some variants of myopic policy (death-weighted myopic, two-day myopic, and seven-day myopic), and shows that the two-day myopic policy is better.

This paper is related to papers discussing equity of disease intervention resources. It has been observed that a more efficient allocation policy typically has a less degree of fairness and vice versa (Kaplan and Merson 2002, Yi and Marathe 2015). Therefore, there are some discussions on the trade-off between efficiency and fairness in the literature. Kaplan and Merson (2002) study the balance between efficiency and equity in allocating HIV-preventing resources. They propose a policy that reserves a proportion of the total resources for fair allocations (allocation proportional to AIDS cases) and uses the remaining resources for cost-efficient allocations. In this paper, we also follow the same idea to balance the efficiency and fairness. Teytelman and Larson (2013) investigate the performance of the convex combination of an efficient switching allocation policy and a pro rata policy. They find that the total number of infections is a decreasing convex function of the weight assigned for the switching allocation policy by numerical experiments. Yi and Marathe (2015) propose a framework to measure fairness of an allocation and use an agent-based SEIR model to derive the relationship between efficiency and degree of fairness under different efficiency measures and fairness axioms. All the above papers observe that a small sacrifice of fairness can have a big decrease in infections. This is also observed in our numerical studies. For other discussion on fairness of allocation, we refer to Lawrence O. Gostin (2009), Huang et al. (2017), and Enayati and Ozaltin (2020).

## 3. Model

In this section, we present an age-structured SAPHIRE model. In this model, the total population is divided into seven compartments including susceptible compartment, exposed compartment, presymptomatic infectious compartment, unascertained infectious compartment, ascertained infectious compartment, isolated compartment, and removed compartment. Each compartment is further divided into five age group (labeled 1,…,5 in ascending order): 0-17, 18-44, 45-64, 65-74, 75+. The dynamics of the age-structured SAPHIRE model is given by the following equations (1)-(7). Figure 1 illustrates the status transition of age group 1 and how susceptible individuals in age group 1 are infected. We should mention that for simplicity, this figure does not specify all the transitions.

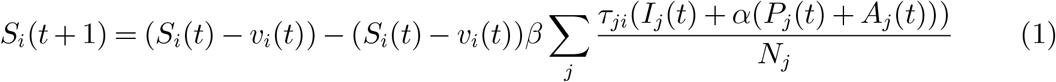

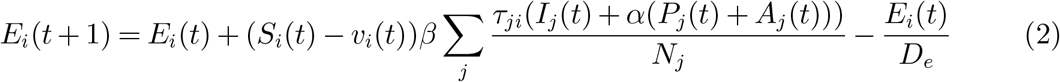

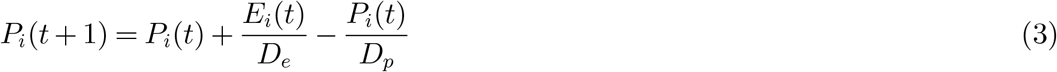

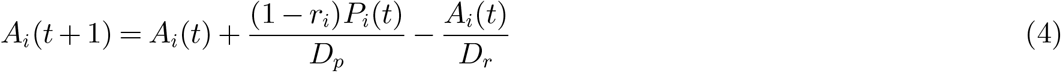

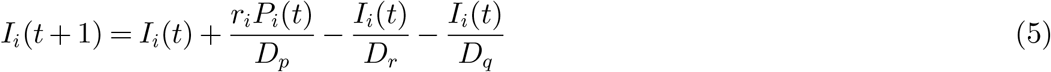

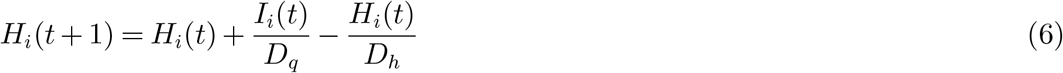

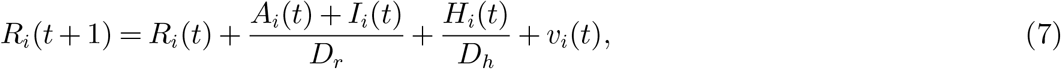

where the meaning of the notations in (1)-(7) are listed in Table 1.

**Figure 1.**
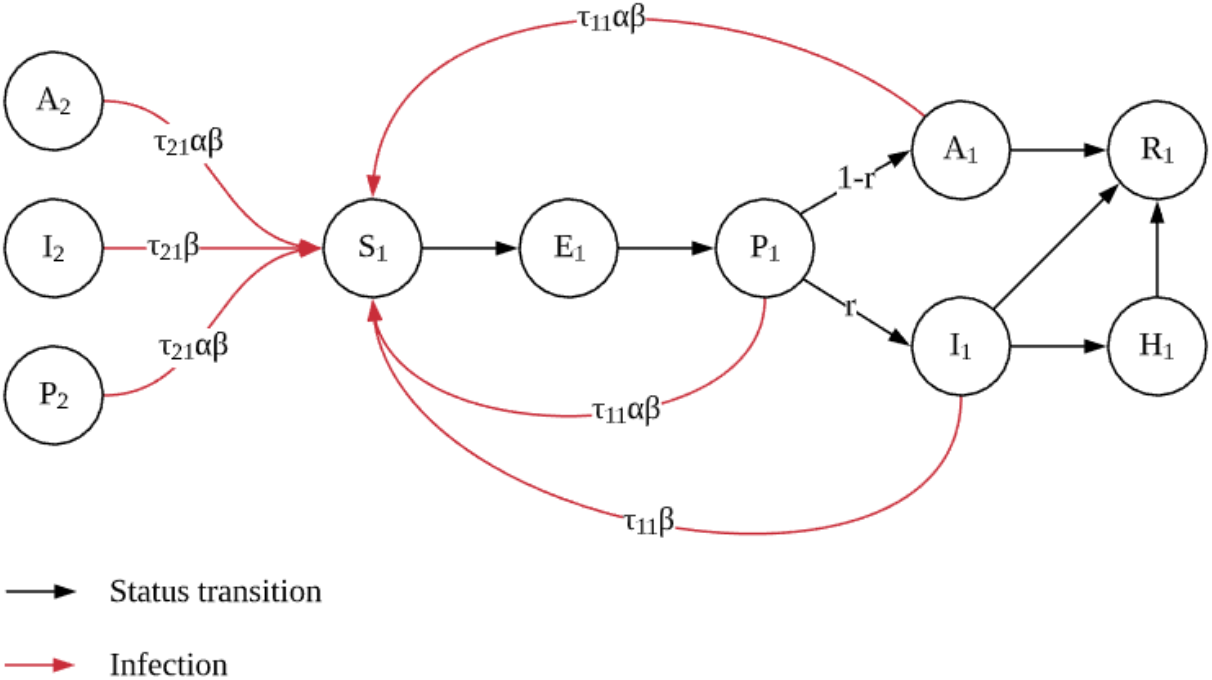
Disease dynamics for the SAPHIRE model.

**Table 1.** Description of notations

|  |  |
| --- | --- |
| $S_i(t)$: | the number of susceptible individuals in age group $i$ at time $t$; |
| $E_i(t)$: | the number of exposed individuals in age group $i$ at time $t$; |
| $P_i(t)$: | the number of presymptomatic infectious individuals in age group $i$ at time $t$; |
| $A_i(t)$: | the number of unascertained infectious individuals in age group $i$ at time $t$; |
| $I_i(t)$: | the number of ascertained infectious individuals in age group $i$ at time $t$; |
| $H_i(t)$: | the number of isolated individuals in age group $i$ at time $t$; |
| $R_i(t)$: | the number of removed individuals in age group $i$ at time $t$; |
| $N_i$: | the population size of age group $i$; |
| $\beta$: | the transmission rate due to the contact between an infectious individual and a susceptible individual; |
| $\tau_{ij}$: | the contact rate of an individual in age group $i$ with an individual in age group $j$; |
| $\alpha$: | the discount factor of the transmission rate due to the contact between an unascertained infectious individual and a susceptible individual; |
| $D_e$: | the average time from exposed to infectious; |
| $r_i$: | the fraction of ascertainment in age group $i$; |
| $D_p$: | the average time from presymptomatic infectious to symptomatic infectious; |
| $D_r$: | the average time from symptomatic infectious to recovered; |
| $D_q$: | the average time from ascertained infectious to isolation; |
| $D_h$: | the average time from isolation to recovered; |
| $v_i(t)$: | the quantity of vaccines allocated to age group $i$ at time $t$. |

In equation (1), *S_i_*(*t*) − *v_j_*(*t*) is the unvaccinated individuals in group *i*, and 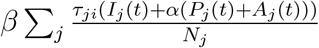 is the force of infection for group *i*, i.e., the rate that an unvaccinated susceptible individual in group *i* gets infected. Here, we assume that only susceptible individuals get vaccination, which can be achieved, e.g., by performing a test prior to vaccination (similar assumption can be found in Lee et al. 2015). For simplicity, we assume that each individual only need one dose of vaccines for immunity and will immediately become immune to the disease after vaccination. This assumption will not affect the qualitative characterization of our allocation policies. Of course, it is more realistic to model the varying probability of being infected after vaccination at different time before immunity is fully established and consider two or more doses vaccination requirement. The transmission of unascertained infectious is discounted by a factor *α* (Li et al. 2020). We use *(τ_ji_)_i,j_* to model the mixing rate of different age groups. See Fumanelli et al. (2012), Liu et al. (2020a) for more on age-specific social contact characterizations. Equation (2) means that the increment of exposed individuals for group *i* equals the new infected individuals in group *i* minus the average number of exposed individuals in group *i* who become presymptomatic (i.e.,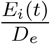). Equation (3) means that the increment of presymptomatic individuals for group *i* equals 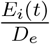 minus the average number of presymptomatic individuals in group *i* who become unascertained infectious (with probability 1 − *r_i_*) or ascertained infectious (with probability *r_i_*). Equation (4) means that the increment of unascertained individuals for group *i* equals the new unascertained infectious individuals transferred from presymptomatic individuals minus the average number of unascertained individuals in group *i* who are removed from the system (recovered and immune to the disease or deceased). Equations (5)(6)(7) have similar explanations. We do not consider birth rate and death rate in our model as the data we use ranges around 3 months and the birth and the death from natural causes can be ignored.

## 4. Data Description and Parameter Estimation

In this section, we describe the data set and how we use it to estimate parameters in our model. The epidemic data we use is disclosed from the NYC Department of Health and Mental Hygiene (NYC Health 2020), which covers the epidemic trajectory of New York City, including daily confirmed cases and deaths, etc. Reported cases are divided by five age groups, 0-17, 18-44, 45-64, 65-74, and 75+ respectively. The age group population information is drawn from a 2017 census (Baruch College 2017). Some summary statistics of different age groups are provided in Table 2. The table includes the total number of confirmed cases and deaths in NYC from March 17 to June 8, as well as the fatality rate of each group. It also covers the proportion of the population that belongs to the group, where the total population in NYC (Manhattan area) is around 1.58 million.

**Table 2.**
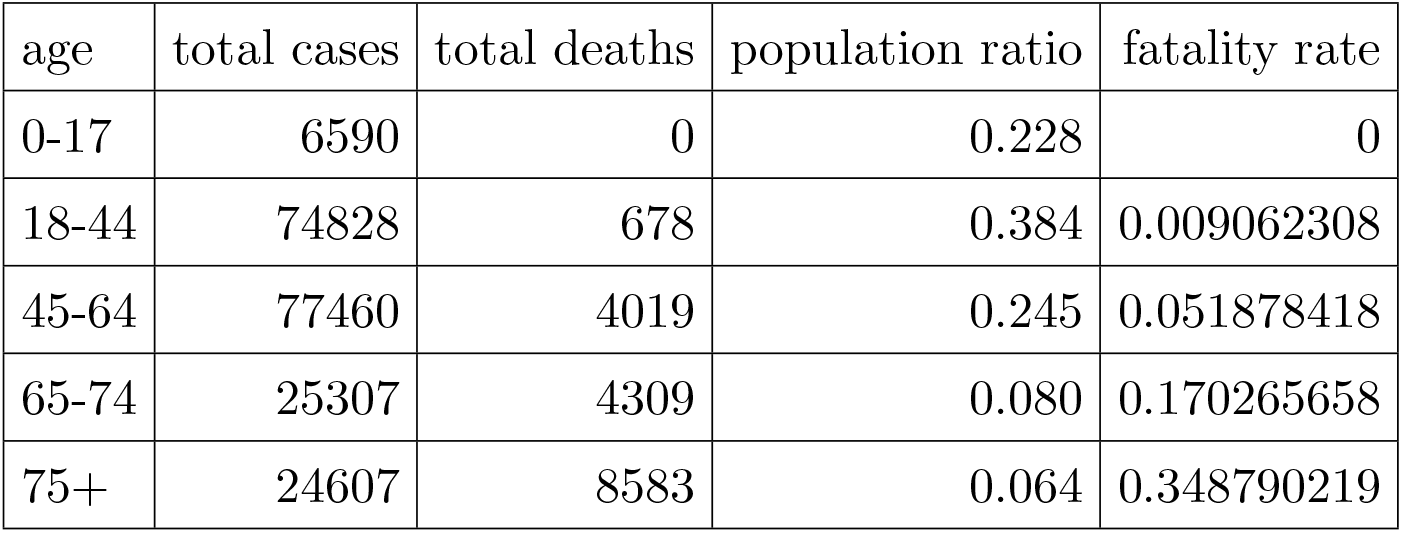
Summary statistics

| age | total cases | total deaths | population ratio | fatality rate |
| --- | --- | --- | --- | --- |
| 0-17 | 6590 | 0 | 0.228 | 0 |
| 18-44 | 74828 | 678 | 0.384 | 0.009062308 |
| 45-64 | 77460 | 4019 | 0.245 | 0.051878418 |
| 65-74 | 25307 | 4309 | 0.080 | 0.170265658 |
| 75+ | 24607 | 8583 | 0.064 | 0.348790219 |

A brief timeline of the epidemic progression and the city’s response is outlined as follows (Wikipedia contributors 2020). On March 1, the first case of COVID-19 was confirmed in New York State. On March 3, the first recorded person-to-person spread cases were confirmed. The epidemic then went through exponential growth, with a number of confirmed cases of 17,800 by March 25. On the other hand, on March 14, all New York public libraries were shut down. On March 17, facilities including theaters, concert venues, and nightclubs were closed. And on March 22, a stay at home order was put into effect where the majority of businesses were paused. As the situation started to turn better in May and June, the city resumed operations according to a four-phase reopening plan, which began on June 8, and on July 20, the final phase was being executed.

We use the epidemic data of New York City, from March 17 to June 8, a total of 84 days, to estimate our model parameters. The reasons for this selection can be summarized as follows. First, New York City is the first major US city struck by the epidemic, and it went through both the increasing phase and declining phase of the epidemic, which provides us rich data about different stages of the epidemic. Second, the time period of our study is approximately during the execution of the stay-at-home order, where the level of social and economical activity stays roughly constant. This simplifies the model as we do not need to incorporate different levels of economic activity in the model, which is not our focus in this paper. Also, since lock-down is a common action taken when facing COVID-19, the estimated model parameters and insights drawn from the model can be more useful in places out of New York City at different times. Third, since we are focusing on the disease spread between different age groups rather than geographical regions, New York city is relatively the best fit because it has a high population density in a rather small area, compared to places like Florida or Texas. But we may expect that the transition parameters among age groups can be quite different at different locations, due to factors including local age structure, level of urbanization, etc.

The way we estimate the parameters is described as follows. Some parameters related to the COVID-19 disease are set according to Hao et al. (2020): *D_r_* = 2.9, *D_h_* = 30, *D_e_* = 2.9, *D_p_* = 2.3, *D_q_* = 6, *β* = 1.4 and *α* = 0.55. Although these parameters are estimated from the epidemic data of Wuhan, we would expect disease-specific parameters to be similar in NYC. See Birge et al. (2020) for the same treatment of using disease-specific parameters from Wuhan for epidemic models of NYC. But we caution that the parameters related to mixing patterns among age groups can be quite different at different locations, this is possibly because of different age structures, different levels of social distancing policies, etc. The ascertainment rates also depend on locations. Hence, we have to estimate the contact rate matrix *(T_ij_*)*_i_*_≤_*_i,j_*_≤5_ among age groups and the ascertainment rate vi,…,v_5_ for the five age groups.

Given a set of all the parameters of the model and the initial number of individuals in each age group and compartment pair, we are able to compute the number of individuals in each age group and compartment pair in all future dates including the daily number of confirmed cases and deaths of each age group. Our objective function (a function of the parameters) is the weighted sum of squared error of the daily number of confirmed cases and deaths predicted by the model to the actual data. The weights are set so that our prediction for confirmed cases and deaths for each age group have similar normalized error. We optimize the objective function with python’s built-in minimization function (SLSQP algorithm). In fact, since the equations that define our model are nonconvex, there exist many local optimal solutions to the objective function and many of them are quite different from each other. In order to address this problem, we impose several inequalities of entries of the contact matrix (revealed from the contact matrix estimated from other papers, e.g., Fumanelli et al. 2012) into the optimization problem. After we incorporate these constraints, the resulting two local optimal solutions that have the lowest objective value are very close to each other with an average percentage difference of 0.375% and a max percentage difference of 4.75%. See Appendix EC.2 for more details. Table 3 presents one of the estimated contact matrices and Table 4 presents the estimated ascertainment rates.

**Table 3.**
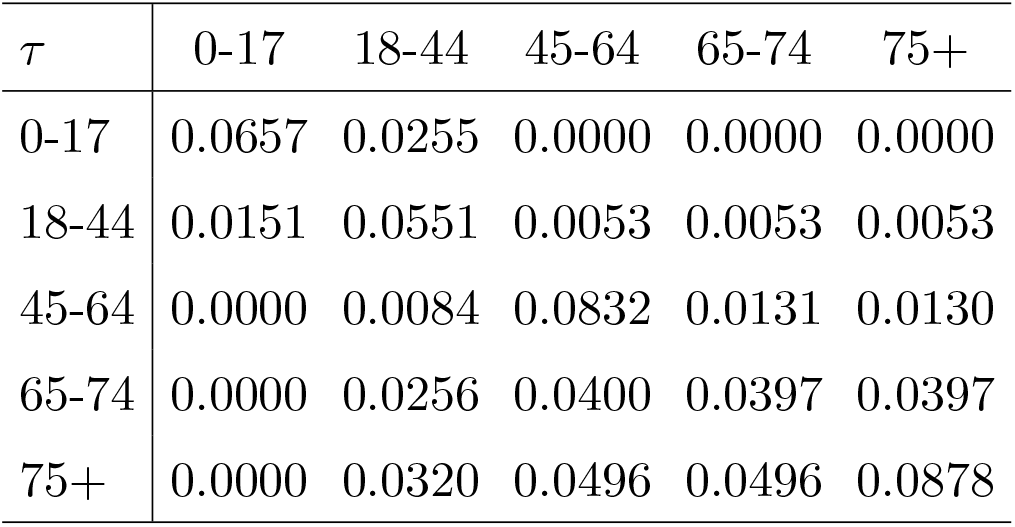
Estimation of contact rates among age groups

**Table 4.** Estimation of ascertainment rates

|  | 0-17 | 18-44 | 45-64 | 65-74 | 75+ |
| --- | --- | --- | --- | --- | --- |
| $r$ | 0.014 | 0.105 | 0.142 | 0.142 | 0.183 |

The plot for the projected confirmed cases against actual data is shown in Figure 2a. Figure 2b shows the projected deaths against actual data. Figure 3a and Firgure 3b represent the projected and actual daily confirmed cases for each age group, respectively. This set of parameters provides a fairly good projection of the confirmed cases and the order of epidemic peaks of all the groups matches the real data. The prediction of the deaths is slightly underestimated with a root mean squared error (RMSE) of 83. However, the accumulated deaths for each group of our model is very close that of the real data. We can see from Figure 4b that the ratio of the accumulated deaths of each age group converges quickly to that of the actual data, and Figure 4a shows the same result.

**Figure 2.**
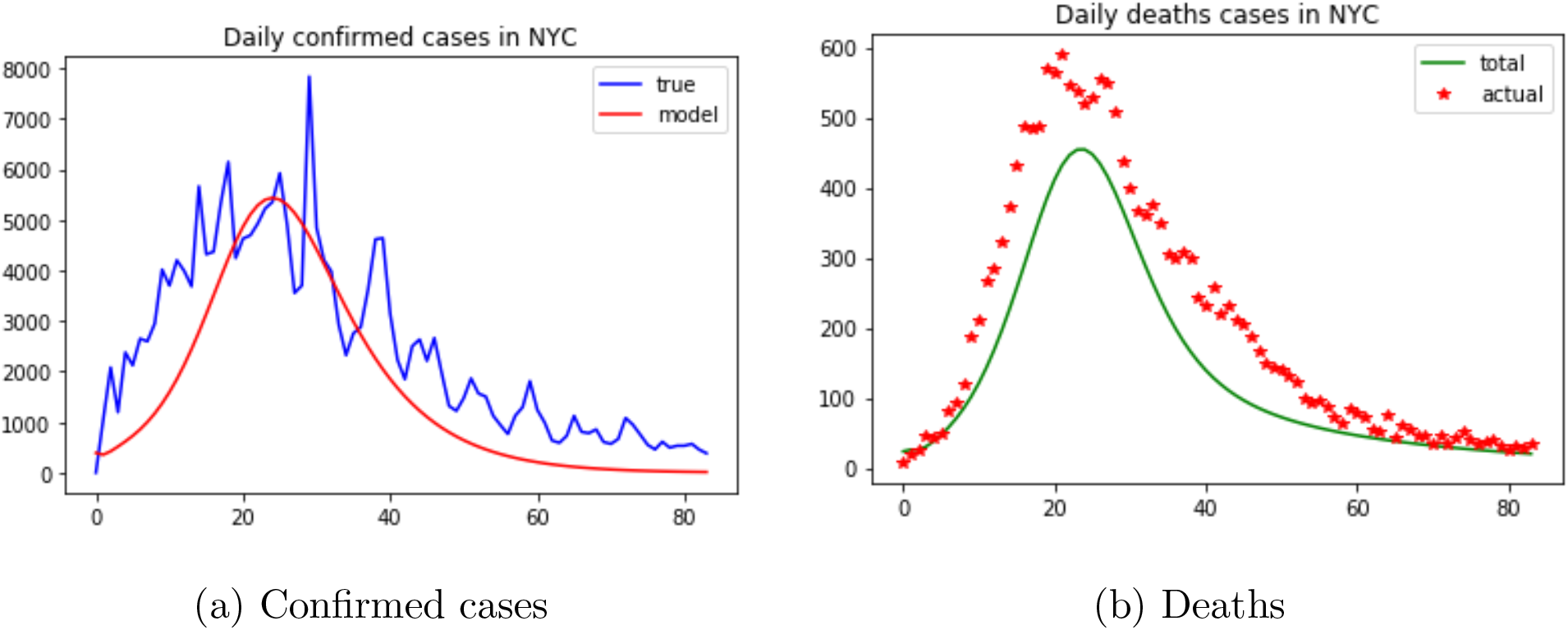
Projected confirmed cases and deaths vs. actual data.

**Figure 3.**
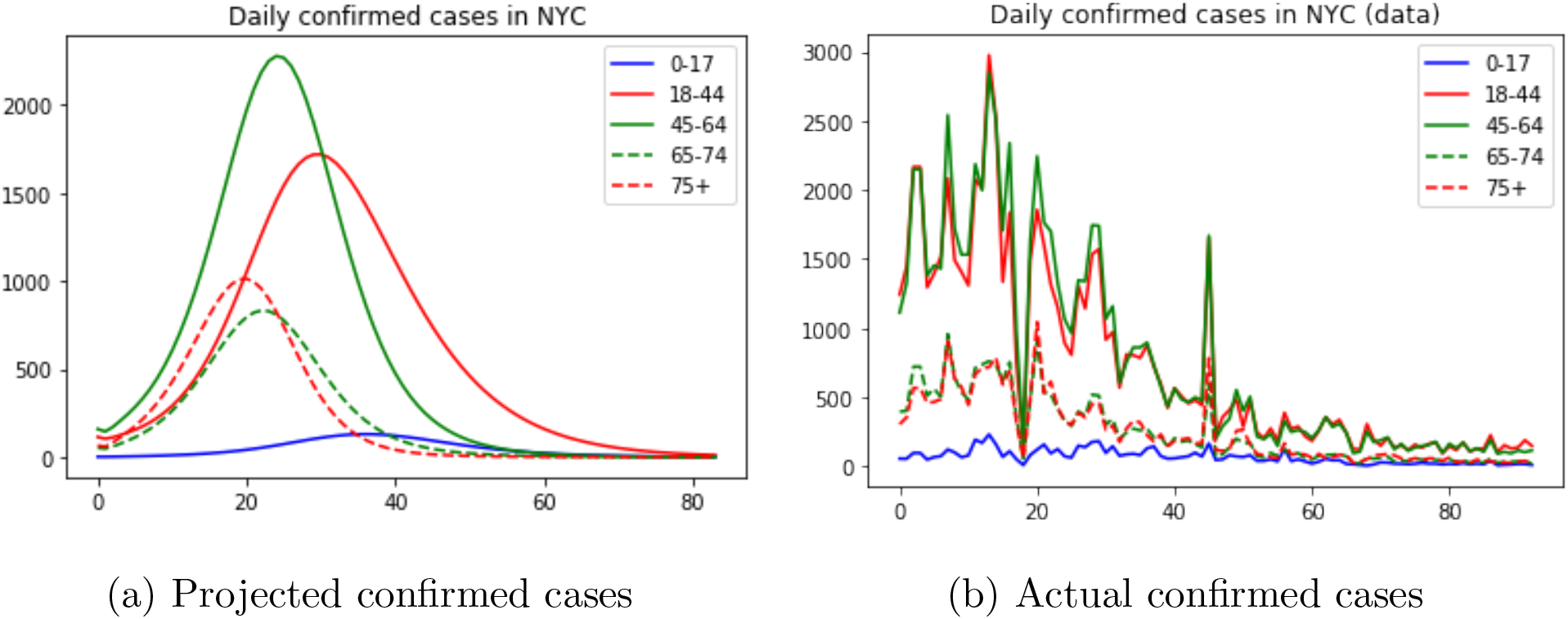
Projected confirmed cases in each age group vs. actual data

**Figure 4.**
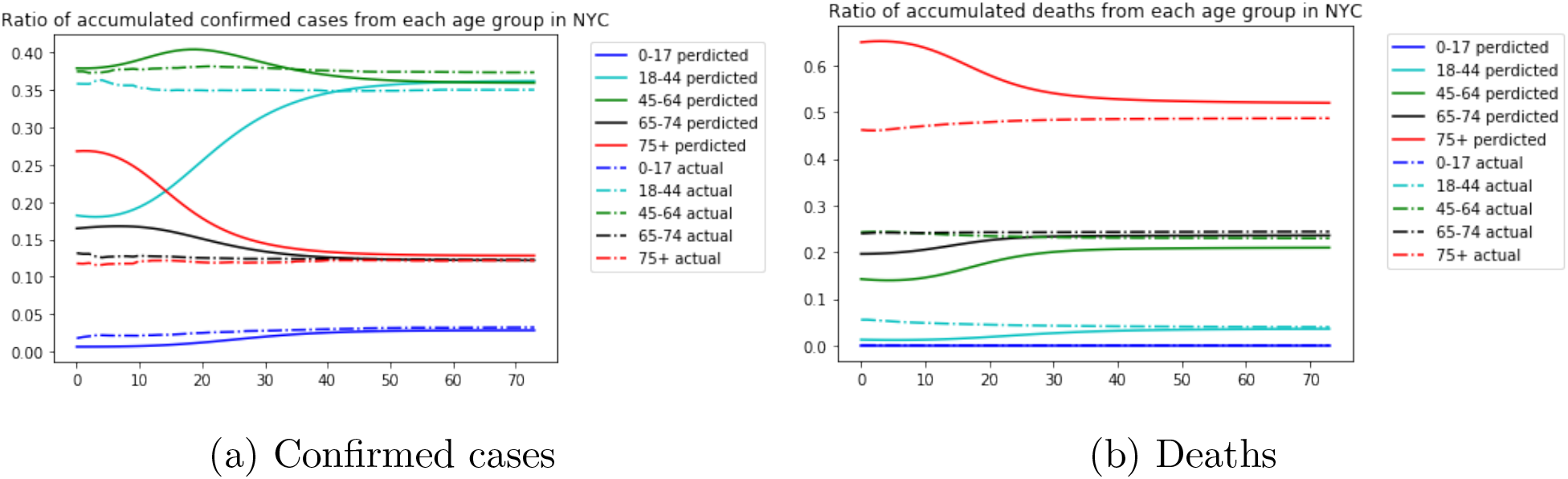
Ratio of accumulated confirmed cases and deaths from each age group in NYC

Some insights can be drawn from the estimated parameters. First, from the scales of *T* we can see that the contact rates are quite heterogeneous. The highest frequency of contact is within the 45-64 group and the 75+ group, which we argue could be because the 45-64 aged people are still carrying out some economical activities during lockdown that are necessary to keep the city running, while for younger people, there are more proportion of office workers who could possibly work remotely and have a smaller frequency of contact with others. For the older group, the 65-74 aged may have a larger proportion of living alone, while the 75+ group may live in nursing homes and contact more often with others. Second, the ascertainment rate for different age groups also varies significantly, with an overall ascertainment rate of around 11%. For the kids, it is actually very small, which partially agrees with the recent news (see, e.g., CBS News 2020) that many children are tested positive on the virus, and would be otherwise not found infected if not tested. This raises a concern that children may also be impacted by the virus. For other groups, it generally follows the order of age, where the older, the more likely to be ascertained. This observation is also consistent with our knowledge of the coronavirus, that it is more dangerous to older people.

## 5. Vaccine Allocation Policy

In this section, we evaluate several vaccine allocation policies (static and dynamic) using our age-structured SAPHIRE model and the parameters estimated. We consider the population of NYC (Manhattan area) in the planning horizon from Mar 17 to June 8 (84 days). Both the case of constant daily supply and the case of increasing daily supply are investigated. We also discuss the trade-off between efficiency and fairness.

### 5.1. Static Allocation

We consider static allocation policies in this subsection, i.e., the proportion of daily available vaccines allocated to each group is the same for each day. Assume that if all the susceptible individuals in a group are vaccinated and there are doses allocated to this group in the future, then the vaccines cannot be transferred to other groups and are wasted. In the following, we derive optimal static allocation policies with respect to different objectives (total confirmed cases, the total deaths, and their weighted summation) under different daily available doses.

#### Constant daily supply case

We assume the daily number of available doses takes values from 2,500 to 15,000. The total doses over the planning horizon will be able to cover from 15% to 80% of the total population. Note that 80% coverage of the total population is sufficient for herd immunity even with a high basic reproduction number of *R*_0_ =3.54 (Hao et al. 2020).

We first consider minimizing the total number of deaths across all groups. For this objective, the vaccine allocation is shown in Table 5. From this table, we can see that when the daily available vaccines are very limited (e.g., 2500 doses), the optimal static allocation policy only focuses on the 75+ age group since they are the most vulnerable to the virus. As the daily available vaccines increase, the policy allocates some doses for younger groups as well. Although these groups are more resistant to severe disease outcomes, providing vaccination to them can help protect the most vulnerable group as well, as they have a large contact rate with the oldest group. Meanwhile, for people aged between 0-44, the policy suggests not to provide vaccination even if the daily available doses increase to 15000. This is because the fatality rates among these groups are extremely low, and their contact levels with the most vulnerable groups are relatively small, so in order to achieve the minimum deaths, we prefer not to give vaccination to them.

**Table 5.** Daily vaccine allocation for minimizing deaths

| doses | 0-17 | 18-44 | 45-64 | 65-74 | 75+ |
| --- | --- | --- | --- | --- | --- |
| 2500 | 0 | 0 | 0 | 0 | 2500 |
| 5000 | 0 | 0 | 0 | 1184 | 3816 |
| 7500 | 0 | 0 | 0 | 2507 | 4993 |
| 10000 | 0 | 0 | 620 | 3322 | 6057 |
| 12500 | 0 | 0 | 2041 | 3779 | 6680 |
| 15000 | 0 | 0 | 3506 | 4102 | 7392 |

We also look at the allocation amount of each age group scaled by their population. Table 6 shows the daily number of vaccine per 10,000 people, allocated to each age group. From this table we can observe that even though the absolute number of vaccines per capita is increased, the scale of increase for younger groups is not that significant as indicated by the previous table. The oldest group still has the largest vaccines per capita when the daily supply of vaccines increases.

**Table 6.** Daily vaccine allocation for minimizing deaths, per 10,000 people

| doses | 0-17 | 18-44 | 45-64 | 65-74 | 75+ |
| --- | --- | --- | --- | --- | --- |
| 2500 | 0 | 0 | 0 | 0 | 246 |
| 5000 | 0 | 0 | 0 | 93 | 376 |
| 7500 | 0 | 0 | 0 | 198 | 492 |
| 10000 | 0 | 0 | 16 | 262 | 597 |
| 12500 | 0 | 0 | 53 | 298 | 658 |
| 15000 | 0 | 0 | 90 | 323 | 728 |

Figure 5a shows the estimated number of deaths as a function of total available doses. The horizontal axis represents the daily available doses, the vertical axis represents the minimal deaths by implementing the optimal static allocation policy. We can see from this figure that the decrease in deaths becomes less significant when the daily available vaccines become large.

**Figure 5.**
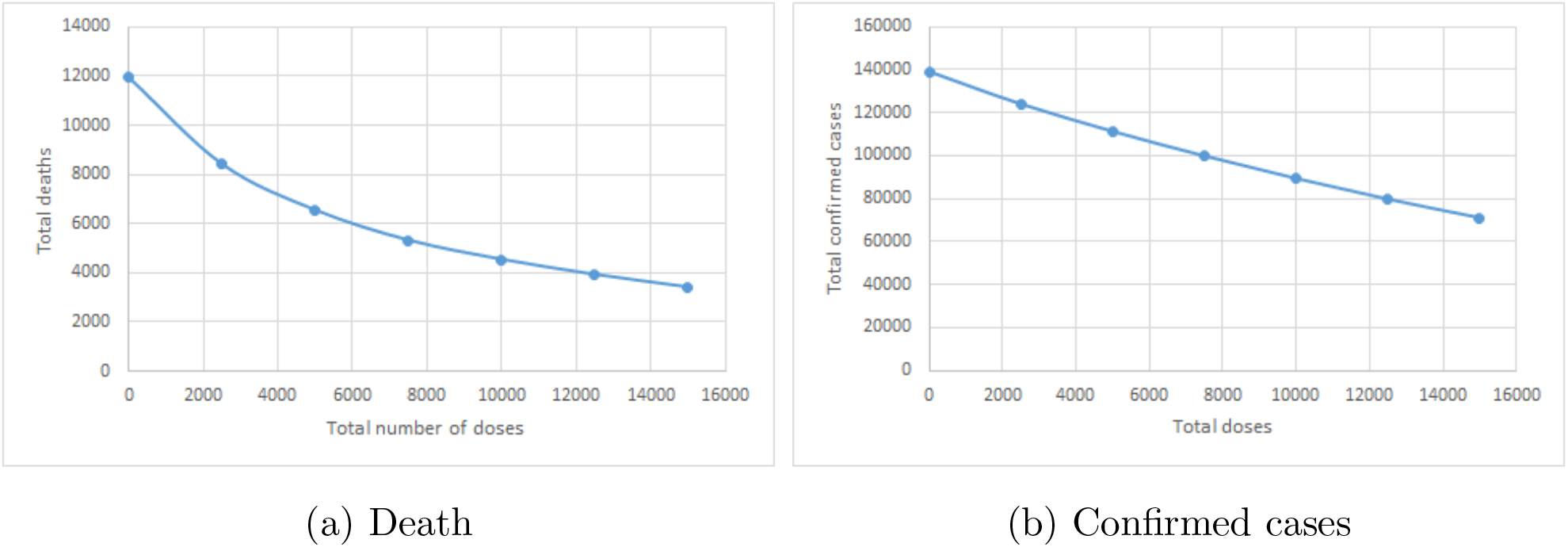
Estimated number of deaths and confirmed cases vs. available doses

We now consider the optimal static policy by minimizing the total confirmed cases from all age groups. Table 7 shows the optimal vaccine allocation to each age group for different daily available doses. We observe from this table that the pattern is similar but Table 5 allocates more to older groups. Besides, the optimal static policy allocates vaccines to younger groups even when the supply is limited. This illustrates that when supply is limited, allocates all the available vaccines to the oldest group has a smaller marginal effect.

**Table 7.**
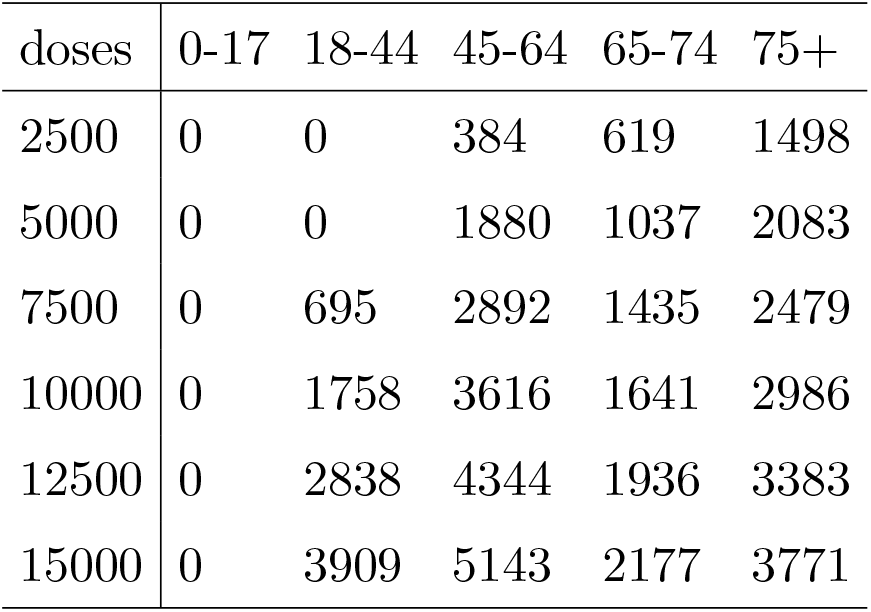
Daily vaccine allocation for minimizing confirmed cases

Again, an allocation per capita and the effect of total doses on total confirmed cases are provided in Table 8 and Figure 5b respectively. We can see that to better prevent the virus from spreading in the population, the oldest group is given less vaccine compared to the previous allocation policy, while other groups are given more to decrease the spread within them. The effect of available doses on the total number of confirmed cases is almost linear, meaning that the incremental doses have equal margins. This indicates that in order to prevent the spread of the virus, it is better to provide as many doses of vaccine as possible, as the increased doses always have a nearly constant marginal effect.

**Table 8.**
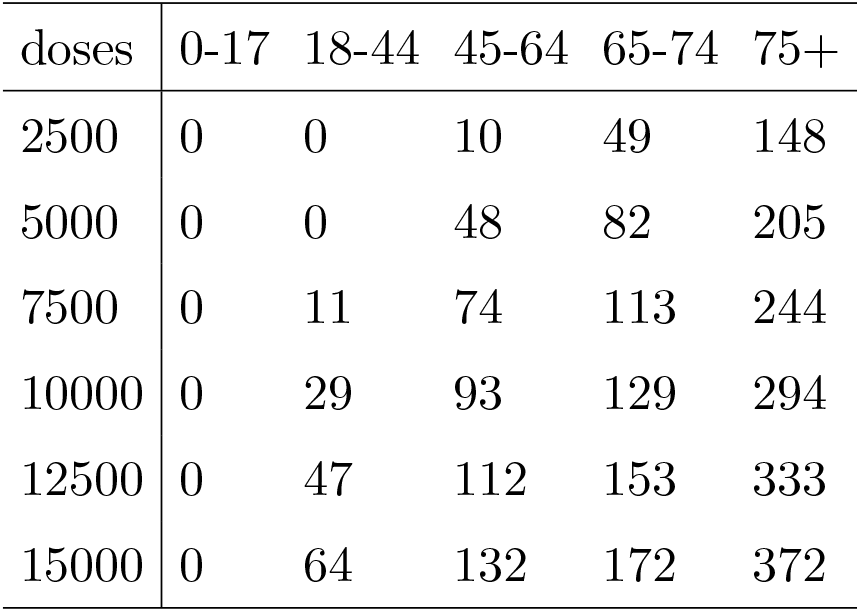
Daily vaccine allocation for minimizing confirmed cases, per 10,000 people

The decision-maker may not simply want to minimize the total confirmed cases or the total deaths. Instead, it is more likely to set an objective that combines these two measures. So, here we consider an objective which is defined as:

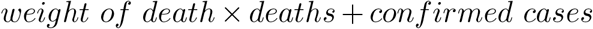

In this case, we fix the total number of daily available doses to be 10,000. Then we increase the weight of death from 1 to 40 to see how the policy is changed. Table 9 shows the optimal allocation under different weight of deaths. We can observe that when death gets more and more weighted, the policy converges slowly to the allocation which minimizes only death and allocates more and more vaccine to the old people. We can see that as we increase the weight of death in our objective, the predicted confirmed cases increase almost linearly, but the decrease of total death becomes very flat quickly. So when making allocation decisions, the decision-maker should properly balance the two objectives, and sometimes the overemphasis of deaths will significantly increase the total confirmed cases, which is not desired as we aim to eventually stop the virus from spreading.

**Table 9.** Daily vaccine allocation for minimizing combined objectives and the outcomes

| death weight | 0-17 | 18-44 | 45-64 | 65-74 | 75+ | total confirmed | total death |
| --- | --- | --- | --- | --- | --- | --- | --- |
| 1 | 0 | 1445 | 3382 | 1786 | 3386 | 89339 | 5619 |
| 5 | 0 | 0 | 3407 | 2451 | 4142 | 90993 | 4915 |
| 10 | 0 | 0 | 2757 | 2687 | 4555 | 92027 | 4756 |
| 15 | 0 | 0 | 2277 | 2719 | 5004 | 93196 | 4659 |
| 20 | 0 | 0 | 2111 | 2884 | 5006 | 93523 | 4641 |
| 25 | 0 | 0 | 1947 | 2893 | 5159 | 94019 | 4620 |
| 30 | 0 | 0 | 1711 | 2907 | 5383 | 94779 | 4592 |
| 35 | 0 | 0 | 1586 | 2914 | 5500 | 95198 | 4579 |
| 40 | 0 | 0 | 1495 | 3004 | 5501 | 95431 | 4573 |

To illustrate the performance of our static policy, we also run two heuristic policies as benchmarks. In the benchmark test, we set the daily available doses to be 10,000 as the results would not change qualitatively if the amount of daily available doses is different. The first benchmark is a uniform allocation policy which gives 2,000 doses for each group per day. Note that the two oldest groups actually have less population, so this benchmark is indeed in favor of the older groups. The total number of confirmed cases under this policy is 95,171, while the number of total deaths is 7051, which is far inferior to what is achieved by the previous policies. The optimal static policy for death minimization has an estimated 98,678 confirmed cases but only 4,541 deaths. The one for confirmed cases minimization achieves 89,183 confirmed cases and 5,942 deaths. Another benchmark is a policy that allocates vaccines to each age group proportionally to its population. This policy has an estimated confirmed case of 100,215 and 9,432 deaths, which is even worse.

#### Increasing daily supply case

In the following, we relax the assumption that the amount of daily available vaccines does not change over time. It is likely that the supply is increasing at the beginning and after some time periods does not change too much. To model this, we assume the amount of daily available vaccines *C_t_* has the following form:

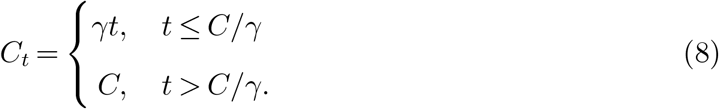

In the simulation study, we set *C* = 10,000, and *γ* takes value between 100 and 600. We consider static policies that allocate a constant percentage of available doses each day to each age group. Under different values of *γ*, the allocation to minimize deaths is shown in Table 10, while the allocation to minimize confirmed cases is in Table 11. We can observe from Table 10 that when the supply is limited at the beginning, i.e., 7 is small, the majority of supplies are allocated to the oldest group. When the supply at the beginning increase, more vaccines are allocated to the group 64-74, but no vaccine is allocated to 0-64 aged individuals. This is almost consistent with the pattern of the optimal static policy when the daily supply is constant. When the objective is minimizing confirmed cases, we observe from Table 11 that groups 18-44 and 45-64 get more percentages of the supply, and their shares do not change too much when 7 varies. This illustrates that when the steady supply *C* is moderately limited (i.e., 10000 in our case), the optimal static policy allocates more to younger groups regardless of the beginning limited supplies.

**Table 10.**
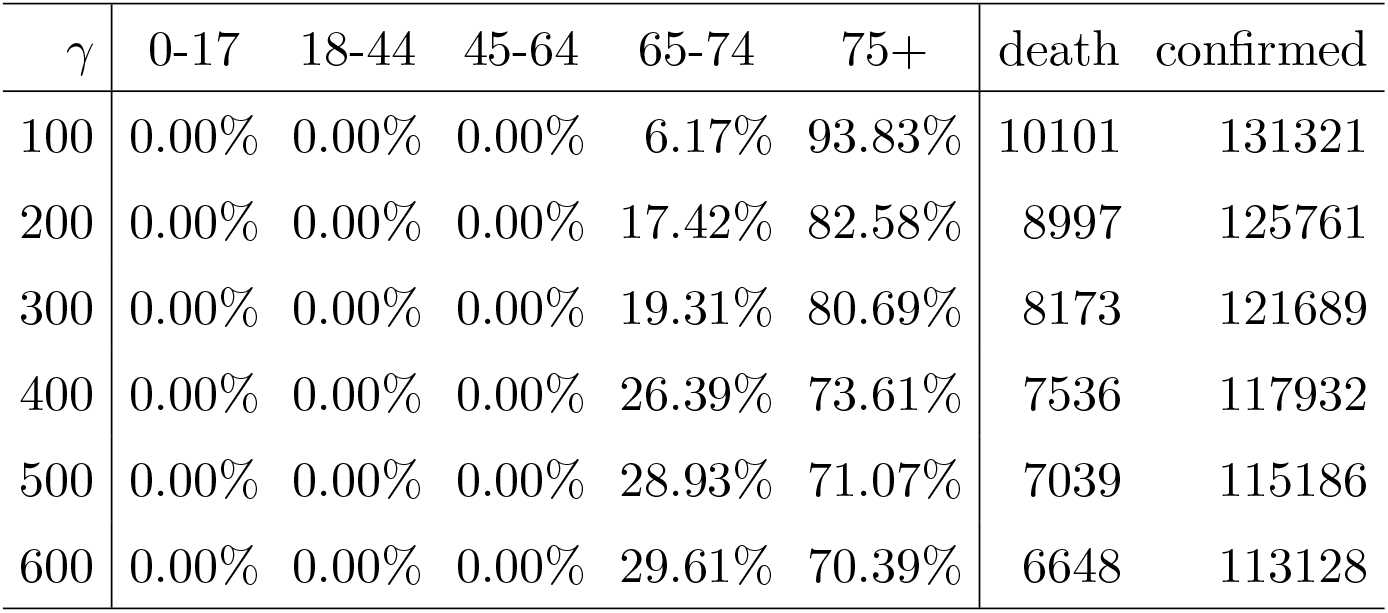
Percentage allocation of minimizing deaths

**Table 11.** Percentage allocation of minimizing total confirmed cases

| $\gamma$ | 0-17 | 18-44 | 45-64 | 65-74 | 75+ | death | confirmed |
| --- | --- | --- | --- | --- | --- | --- | --- |
| 100 | 0.00% | 31.53% | 27.50% | 15.36% | 25.62% | 10946 | 129580 |
| 200 | 0.00% | 37.00% | 29.80% | 12.42% | 20.79% | 10390 | 121859 |
| 300 | 0.00% | 36.21% | 32.63% | 12.07% | 19.11% | 9876 | 115438 |
| 400 | 0.00% | 31.87% | 33.50% | 13.56% | 21.12% | 9238 | 110503 |
| 500 | 0.00% | 28.74% | 34.49% | 13.95% | 22.87% | 8712 | 106860 |
| 600 | 0.00% | 27.24% | 35.58% | 15.24% | 21.99% | 8402 | 104181 |

We mention that under the assumption that superfluous vaccines allocated to a group are wasted, the optimal static policy will cause a huge waste even when the supply is limited. For example, when the daily supply is a constant 2500, the total vaccines allocated to group 75+ is 84 × 2500 = 210000, which is more than twice of its population 101120. This suggests considering dynamic allocation policies which are examined in the next section.

### 5.2. Dynamic Allocation

We consider dynamic allocation policies in this section. Since our age-structured SAPHIRE model has 35 age-compartment pairs and the population in each pair may range to several thousand (some ranges to hundreds of thousands, e.g., susceptible compartment), it is challenging to compute the optimal dynamic allocation using dynamic programming. One may want to use approximate dynamic programming to obtain good heuristics. However, as illustrated in Long et al. (2018) with a space-structured epidemic model for the 2014 Ebola outbreak, where the authors evaluate allocation policies of Ebola treatment beds to different regions (not vaccine allocation to different age groups considered in this paper), the heuristic obtained from approximate dynamic programming performs worse than simple heuristics such as myopic policy (to be discussed below for our model). Hence, we only provide evaluations of several dynamic allocation heuristics.

- (**Old-First Policy**) This policy allocates available daily vaccines to age group 5 first. If there are any remaining vaccines, it allocates to age group 4, and so on.
- (**Infection-First Policy**) This policy allocates available daily vaccines proportionally to the infection ratio of each group (i.e., 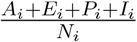).
- (**Myopic Policy**) In each time *t*, the myopic policy determines the amount of vaccines *v_i_* allocated to age group *i* by minimizing the total new infections of all groups in time *t* + 1, i.e.,

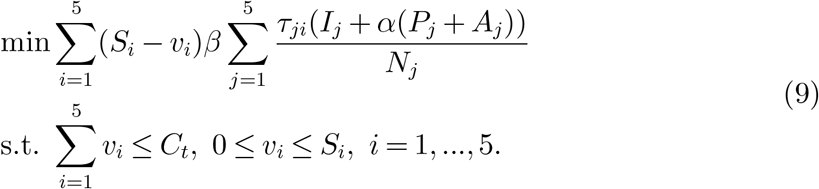 Since this is a linear program with a capacity constraint and a box constraint, the myopic policy will first allocate vaccines to the group with the largest coefficient. If the number of susceptible individuals in this group is less than *C_t_*, then the myopic policy allocates the remaining doses to the group with the second-largest coefficient, and so on. The coefficient of *v_i_* in the objective can be regarded as the marginal effect of a unit vaccine allocated to group *i*.
- (**Death-Weighted Myopic Policy**) In each time *t*, the death-weighted myopic policy determines the number of vaccines *v_i_* allocated to age group *i* by minimizing the total weighted infections of all groups in time *t* + 1,

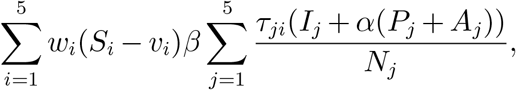

where *w_i_* is the death rate of group *i*.
- (**Two-Day Myopic Policy**) In each time *t* = 1,3,5,…, this policy determines *v_i_*(*t*)*,v_i_(t* + 1), *i* = 1,…,5 to minimize the new infections of all groups in time *t* + 2, i.e.,

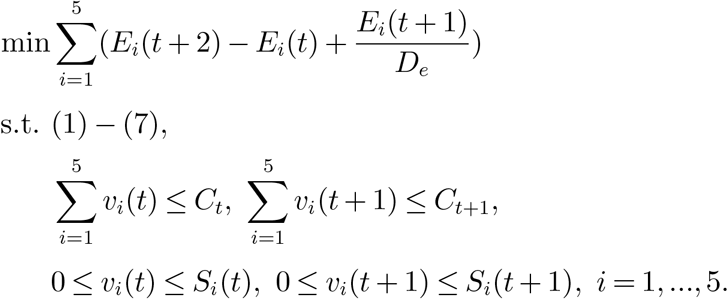 Then the allocations *v_i_*(*t*) and *v_i_(t* +1) are implemented in time *t* and *t* +1, respectively.
- (**Seven-Day Myopic Policy**) In each time *t* = 1,8,15,…, the policy determines *v_i_*(*t* + *j*), *i* = 1,…, 5, *j* = 0,…, 6 to minimize the new infections of all groups in time *t* + 7. Then the allocations *v_i_(t* + *j*), *i* = 1,…, 5, *j* = 0,…, 6 are implemented in time *t*, *t* +1,…, *t* + 6.

#### Constant daily supply case

We set the daily supply *C_t_* to be a constant 10,000. For the two-day myopic policy and seven-day myopic policy, as we mentioned in Section 4, the disease dynamics (1)-(7) is nonlinear. Hence, their corresponding optimization problems are nonconvex problems, which potentially have many local optimal solutions. To obtain good local optimal solutions, we solve the optimization problems 100 times at the beginning of every two days, or seven days and pick the one that gives the minimal objective value. The outcomes of the above heuristics are listed in Table 12.

Here, we do not present the outcome of the death-weighted myopic policy as it yields the same allocation with the old-first policy in this numerical study. Table 12 shows that the two-day myopic policy performs the best in terms of both the total confirmed cases and the total deaths. The myopic policy and the seven-day myopic policy also perform well. The myopic policy has slightly more deaths than the seven-day myopic policy, but the later has slightly more confirmed cases. This demonstrates that myopic policies with longer planning days do not perform better. The old-first policy has the largest total confirmed cases and the infection-first policy has the largest total deaths. This is because the old-first policy postpones younger groups that have the most infections, and the infection-first policy only considers the infection ratio 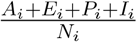. Although the old-first policy itself seems to be beneficial to decrease deaths, it actually causes more deaths compared to the myopic policy and the two-day myopic policy which do not explicitly minimize deaths in their optimization problems. This illustrates that the decreasing of confirmed cases leads to fewer deaths as well and a good allocation can decrease both deaths and confirmed cases.

**Table 12.** Outcomes of different dynamic heuristics

|  | confirmed |  |  |  |  |  | death |  |  |  |  |  |
| --- | --- | --- | --- | --- | --- | --- | --- | --- | --- | --- | --- | --- |
|  | 0-17 | 18-44 | 45-64 | 65-74 | 75+ | total | 0-17 | 18-44 | 45-64 | 65-74 | 75+ | total |
| old first | 3638 | 42787 | 35937 | 5709 | 1898 | 89969 | 0 | 359 | 1792 | 954 | 676 | 3781 |
| myopic | 2107 | 35589 | 21827 | 6185 | 1831 | 67540 | 0 | 296 | 1085 | 1008 | 639 | 3028 |
| infection-first | 2285 | 31254 | 29341 | 5576 | 4315 | 72770 | 0 | 263 | 1450 | 923 | 1482 | 4118 |
| two-day myopic | 2121 | 35382 | 21735 | 6696 | 883 | 66818 | 0 | 295 | 1081 | 1090 | 332 | 2798 |
| seven-day myopic | 2304 | 33098 | 26681 | 4364 | 1581 | 68028 | 0 | 276 | 1320 | 728 | 568 | 2892 |

In the following, we compare the allocation of different dynamic policies. Since the old-first and the myopic policy only allocate the daily supply to one group (except when the remaining susceptible individuals of the group cannot consume all the supply), we can visualize them as in Figure 6. In the figure, the different color blocks represent time periods of the allocation to different groups, and the vertical axis represents the time. The first row corresponds to the old-first policy, and the second row corresponds to the myopic policy. We do not specify the mixing of allocations on the boundary of different color blocks. The coefficients of v_i_ for different age groups under the myopic policy is shown in Figure 7. Note that even though the group 75+ has the largest coefficient from day 10 (starts to allocate to group 65-74) to 35, the vaccines will not be given to group 75+ after day 10 as all its susceptible individuals are vaccinated. The allocation of the two-day myopic policy is shown in Figure 8a.

**Figure 6.**
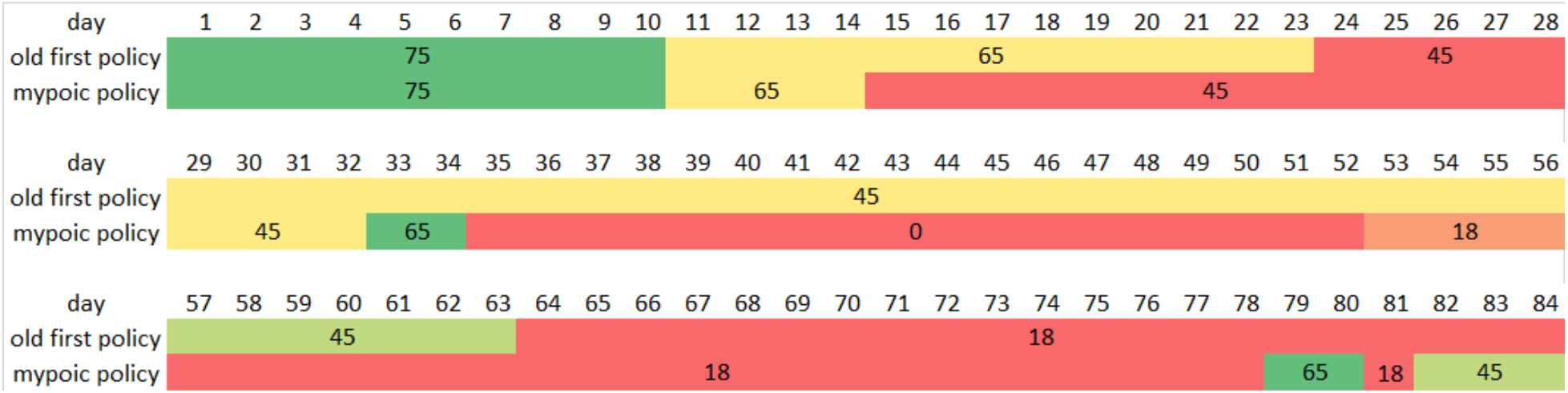
Allocation for old-first policy and myopic policy

We can observe that the execution of the myopic policy follows a similar pattern as the old-first policy at the beginning in that they both start from the oldest age group and gradually move to younger groups. There are some differences between these two policies that explain the superiority of the myopic policy. First, the myopic policy spends less time in the 65-74 and 45-64 age groups. We can observe that the myopic policy does have slightly more confirmed and deaths for the 65-74 group. Although the myopic policy spends less time in these groups, it moves to other groups (0-17, 18-44) with larger vaccine marginal effect (see Figure 7), and helps to control the disease transmission in all groups. Second, the old-first policy still allocates vaccines to group 45-64 after day 34 even though the vaccine marginal effect of this group is smaller than group 0-17, while the myopic policy skips the 18-44 age group at day 34 and goes directly to the 0-17 age group. At the end of the horizon, the myopic policy fluctuates among the groups 65-74, 18-44, and 45-64 as their vaccine marginal effects are nearly the same and short periods of allocation will change the order of vaccine marginal effects of different groups. At this stage, the allocation is insensitive to different groups and a change of the myopic policy will not hurt the performance.

**Figure 7.**
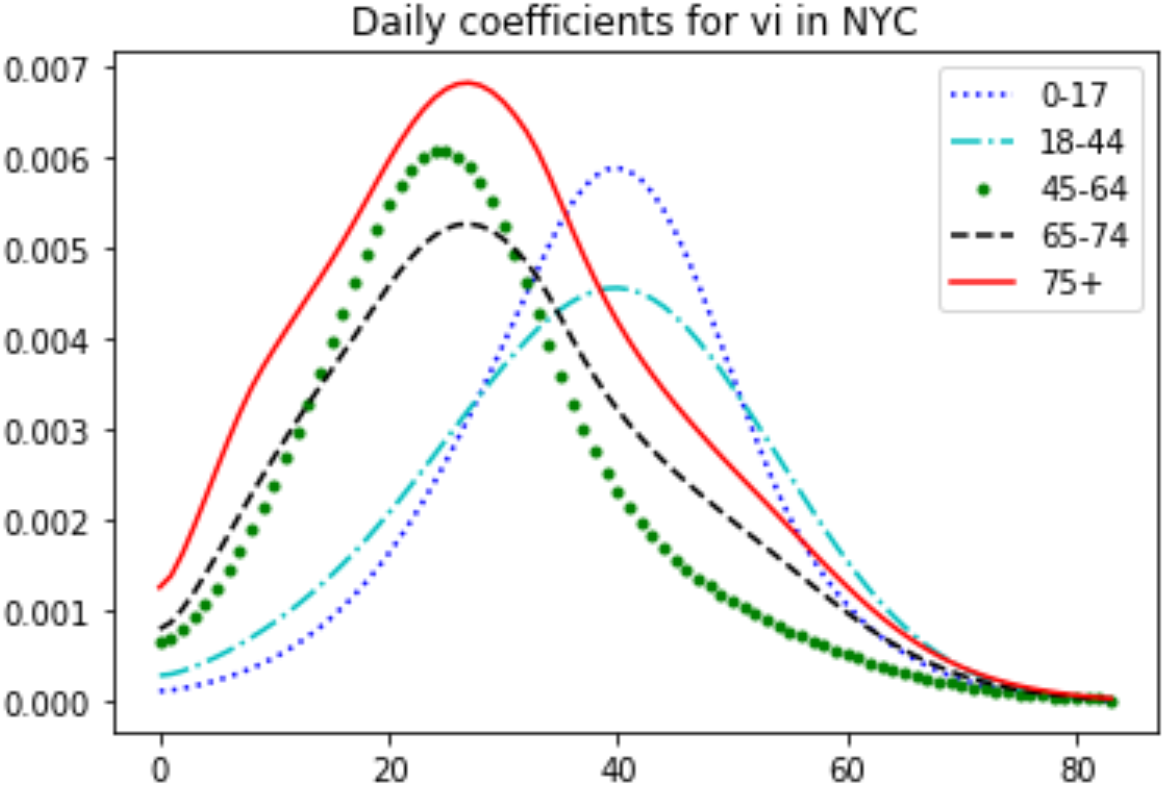
Coefficients of *v_i_* for different age groups under myopic policy

The two-day myopic policy has a mixing allocation as shown in Figure 8a. The majority of the supply is first given to the group 75+ and moves to the group 65-74 roughly on day 10. Around day 15 the majority of the supply is given to 45-64 and then moves to the group 0-17 around day 34. Then the policy gives most of the vaccines to the group 18-44 after day 52. Comparing with the allocation of the myopic policy (Figure 6), the two-day myopic policy roughly follows the pattern of the myopic policy but has more mixing on the exchange time boundary and the end of the horizon, which may be the reason for its slightly better outcomes.

**Figure 8.**
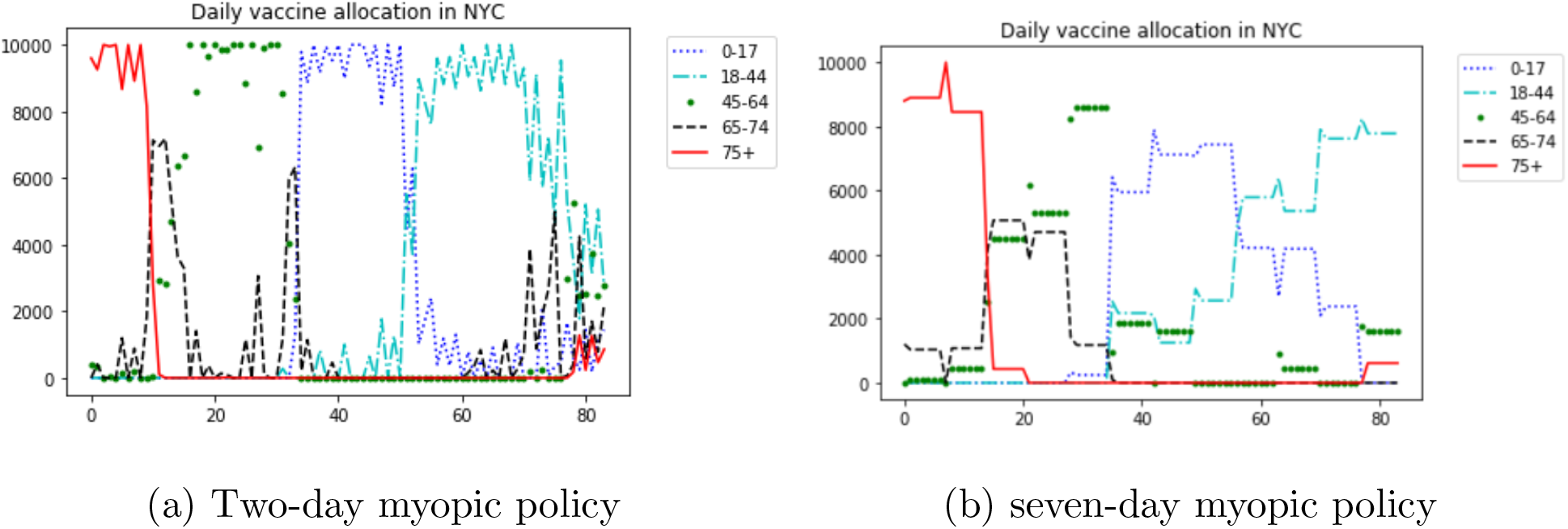
Daily allocation under different myopic policies

The seven-day myopic policy is shown in Figure 8b. As we can see from this figure, the daily allocation of this policy still follows a similar pattern as the two-day myopic policy, but with a higher level of mixing in the allocation.

#### Increasing daily supply case

In addition to the case where the daily supply is constant throughout the time horizon, we also consider the setting that the daily supply is gradually increasing over time. Again, the daily supply follows equation (8), *C* = 10,000, and *γ* take values 100, 200,…, 600. The outcomes of the two-day myopic policy, the old-first policy, and the infection-first policy are shown in Table 13. We do not put the outcome of the myopic policy here as it is very close to that of the two-day myopic policy. The outcomes of static policies are put here for comparison convenience.

**Table 13.** Performance of dynamic policies under increasing vaccine supply

|  | two-day myopic policy |  | old-first policy |  | infection-first |  | static: confirm |  | static: death |  |
| --- | --- | --- | --- | --- | --- | --- | --- | --- | --- | --- |
| $\gamma$ | confirmed | death | confirmed | death | confirmed | death | confirmed | death | confirmed | death |
| 100 | 123531 | 9208 | 123625 | 9087 | 125350 | 10324 | 129580 | 10946 | 131321 | 10101 |
| 200 | 111794 | 7612 | 111804 | 7488 | 114662 | 9128 | 121859 | 10390 | 125761 | 8997 |
| 300 | 102456 | 6515 | 101729 | 6377 | 105963 | 8143 | 115438 | 9876 | 121689 | 8173 |
| 400 | 95900 | 5671 | 96172 | 5494 | 99628 | 7367 | 110503 | 9238 | 117932 | 7536 |
| 500 | 91361 | 5189 | 93548 | 5269 | 95176 | 6800 | 106860 | 8712 | 115186 | 7039 |
| 600 | 88031 | 4750 | 91677 | 5148 | 91992 | 6396 | 104181 | 8402 | 113128 | 6648 |

From Table 13 we can observe that dynamic policies outperform static policies. Unlike the case where daily supply is constant, the performance of the old-first policy outperforms the infection-first policy and is close to that of the two-day myopic policy. Comparable outcomes of the old-first policy and the two-day myopic policy may due to their similar allocations during the beginning of the planning horizon. However, their performances are all worse than those under constant daily supply. This demonstrates that one may not achieve a significantly better performance when the starting inventory is extremely limited.

The dynamic allocation of the two-day myopic policy for *γ* = 100 is shown in Figure 9. We can see that the allocation follows a similar pattern to the case when vaccine supply is constant over time. However, it spends much more time on the oldest group since the initial available vaccines are more limited. Similar observation can be found under other values of 7.

**Figure 9.**
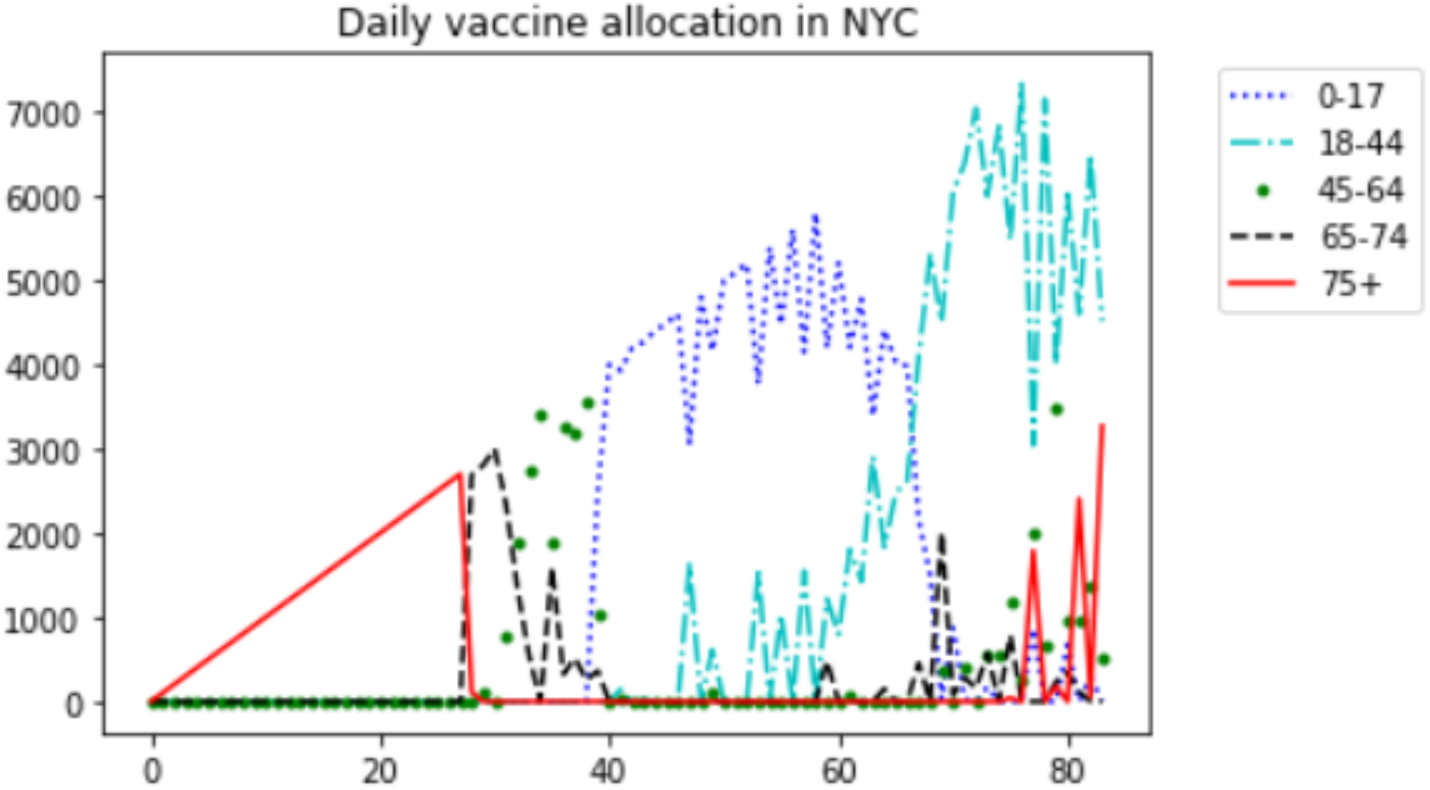
Two-day myopic allocation when *γ* = 100

### 5.3. Impact of fairness

So far, we only look at the efficiency of allocation policies. However, as the ACIP COVID-19 vaccines work group points out, equity in vaccine allocation and distribution is one of the primary goals (Dooling 2020). We thus discuss fairness in this subsection.

In this study, we consider a policy that is a combination of the pro rata policy and the myopic policy to account for the trade-off between equity and efficiency as the pro rata policy is regarded as the fairest policy (implemented by CDC for the H1N1 vaccine allocation to states (Centers for Disease Control and Prevention 2010)) and the myopic policy is one of the most efficient policies discussed in this paper. Let λ ∈ [0,1] be the fraction of daily vaccine doses reserved for efficient allocation. Denote 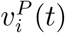 the allocation quantity of the pro rata policy with daily supply (1 − λ)*C_t_* and 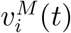 the allocation quantity of the myopic policy with daily supply *λC_t_*. Consider the allocation policy 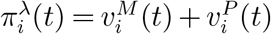. Obviously, this policy degenerates to the myopic policy when *λ* =1, and it degenerates to the pro rata policy when *λ* = 0. Figure 10a and Figure 10b shows the number of total confirmed cases and deaths under different fraction of vaccine doses using myopic policy. The vertical axis reports the total confirmed cases or total deaths, and the horizontal axis reports the value of *λ*. It is clear that the two curves are deceasing in *λ* as more fraction of myopic allocation means more efficiency.

**Figure 10.**
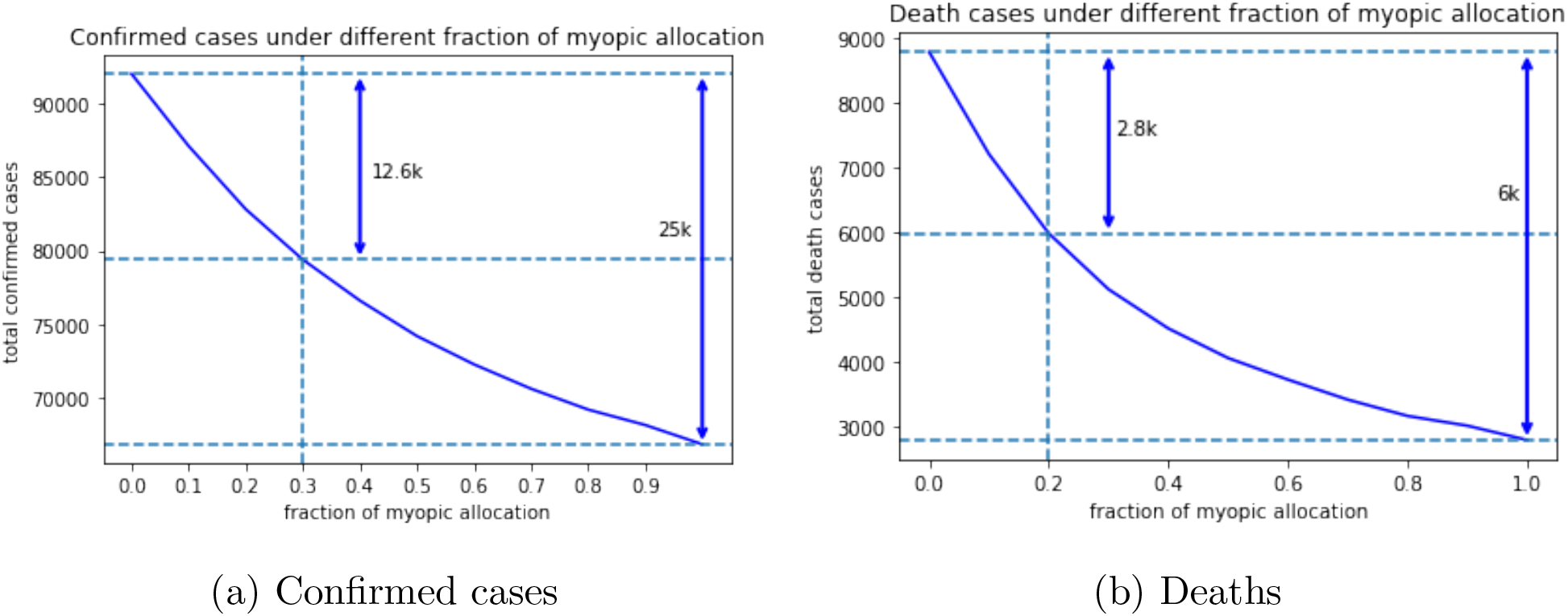
Confirmed cases and deaths under different

Similarly to Kaplan and Merson (2002) and Teytelman and Larson (2013), we find decreasing marginal return of confirmed cases and deaths as we increase the amount of reserved capacity for the myopic policy. For example, Figure 10a depicts the total confirmed cases as a function of capacity reserved for the myopic policy. When no capacity is reserved for the myopic policy, the total confirmed cases is 92,017, while when all capacity is reserved for the myopic policy, the total confirmed cases is 66,815 (roughly 25,000 cases averted). Surprisingly, reserving only 30% of the capacity for the myopic policy reduces the number of confirmed cases from 92,017 to 79,382, a 50% reduction (roughly 12,600 cases) in the averted cases associated with pure myopic policy.

The impact is even stronger when measuring the number of deaths (see Figure 10b). Specifically, in this case allocating only 20% of the capacity to the myopic policy achieves a 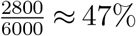 reduction in averted deaths associated with pure myopic policy.

Note that here we measure the fairness by the fraction of the daily capacity reserved for the pro rata policy. This is not the only measure of fairness. In fact, some other fairness measures (e.g., Gini index) are employed to evaluate vaccine allocation policies in the literature (see Yi and Marathe 2015 and Enayati and Ozaltin 2020). See Appendix EC.1 for the impact of fairness measured by Gini index on myopic policies.

## 6. Conclusion

This paper considers vaccine allocation policies of COVID-19 to different age groups under limited supply. We use an age-structured SAPHIRE model and estimate relevant parameters with the epidemic data from NYC. Base on this model and the estimated parameters, we evaluate the performance of the optimal static policies and several dynamic allocation heuristics under different settings of the daily vaccine supply. Our numerical study shows that generally, the optimal static policy allocates most of the vaccines to older groups when the objective is minimizing deaths, and if the objective is minimizing confirmed cases, then younger groups will get more. This suggests the decision-maker needs to balance very carefully between different objectives in order to derive policies that perform much better than ad-hoc allocation policies. The dynamic allocation heuristics in general perform better than the static ones. Among the dynamic policies, the best are myopic policies (including two-day, seven-day), and they perform much better than the other heuristics. We also discuss the trade-off between equity and efficiency. We show that high efficiency can be achieved by sacrificing a small portion of equity.

Our paper though has the following limitations. One is on the assumption placed on static policies, i.e., once the people in one group have all been vaccinated, the doses allocated to that group cannot be transferred to other groups and are wasted. This is not the case in reality, and it would be interesting to see how the static policy performs when the transfer of vaccines to other groups is allowed. Alternatively, a more complicated policy, for instance, piecewise static policies can be used to reduce the waste of vaccines under this assumption. Another limitation is that we do not solve the optimal dynamic policy. This is due to the nonlinearity of the disease dynamics leading to a highly nonconvex optimization problem, which is a subject for future research. Finally, our numerical study only uses data from NYC. It would be interesting to extend our study to incorporate spatial structures. Nevertheless, we believe the insights of allocation policies drawn from it could provide a valuable reference to decision-makers on the allocation of the upcoming COVID-19 vaccines.

## Data Availability

The datasets used in this paper are available in the NYC Department of Health and Mental Hygiene and Baruch College.

https://www1.nyc.gov/site/doh/covid/covid-19-data.page

https://www.baruch.cuny.edu/nycdata/population-geography/age_distribution.htm

## Acknowledgments

The authors are grateful for the valuable suggestions and comments from Prof. Edward H. Kaplan which greatly improve this paper.

## Electronic companion

### EC.1. Impact of Fairness Measured by Gini Index

We employ the concept of Gini index to measure the degree of fairness. Gini index (or Gini coefficient) is a measure of statistical dispersion which is often used to reflect the degree of income inequality. It is also used in Enayati and Özaltin (2020) to measure the inequality of influenza vaccine allocation. Given a nonnegative vector *x* = (*x_i_*,…,*x_m_*), the Gini index of x is defined as

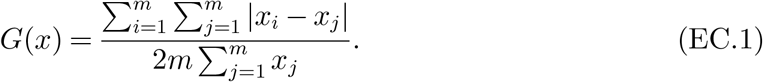

Here, x can represent, for example, the faction of total capacities allocated to group *i*. It is clear that a uniform allocation (i.e., *x*_1_ = … = *x_m_)* which means perfect equality has a Gini index of zero. An allocation with a Gini index of one means maximal inequality. Smaller Gini index means more fairness. In the following, we seek to find efficient allocation policies with small Gini index. To achieve this, we impose an additional constraint to the LP (9) that requires the Gini index of the normalized allocation 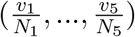 to be no more than ϵ, where ϵ is a tolerance of inequality, as follows.

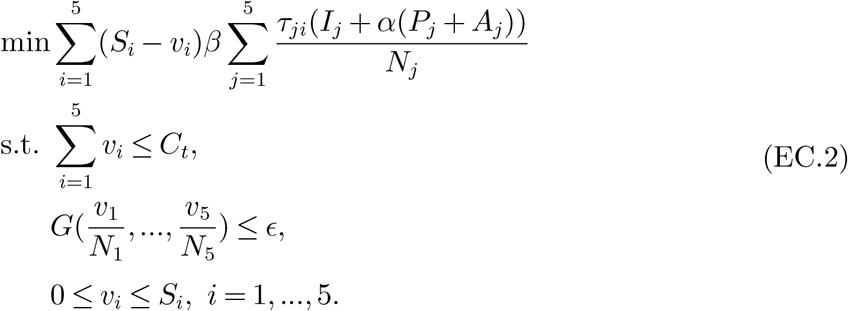

Figure EC.1 and Figure EC.2 show the total number of confirmed cases and deaths achieved by the myopic policy under different level of inequality tolerance ϵ, respectively. We observe that the two curves are almost linear, which means equal marginal return of the confirmed cases and deaths when more inequality is allowed.

Figure EC.3 presents the fraction of total capacities allocated to each group and Figure EC.4 presents the vaccine coverage of each group. We can see that when the degree of fairness increases (i.e., ϵ decreases), the group 18-44 gets more doses while the older groups 65-74, 75+ get fewer doses. This is because the group 18-44 has a very large population (38% of the total population). In terms of the vaccine coverage, when the degree of fairness increases, all the groups converge to the vaccine coverage of 53%. In particular, the vaccine coverage for the older groups are decreasing, the vaccine coverage of the group 18-44 is increasing, and the vaccine coverage of other groups dose not change too much. We caution the reader that it is easy to tell which allocation is fairer by comparing the Gini index. However, the numerical value of Gini index is less interpretable.

**Figure EC.1.**
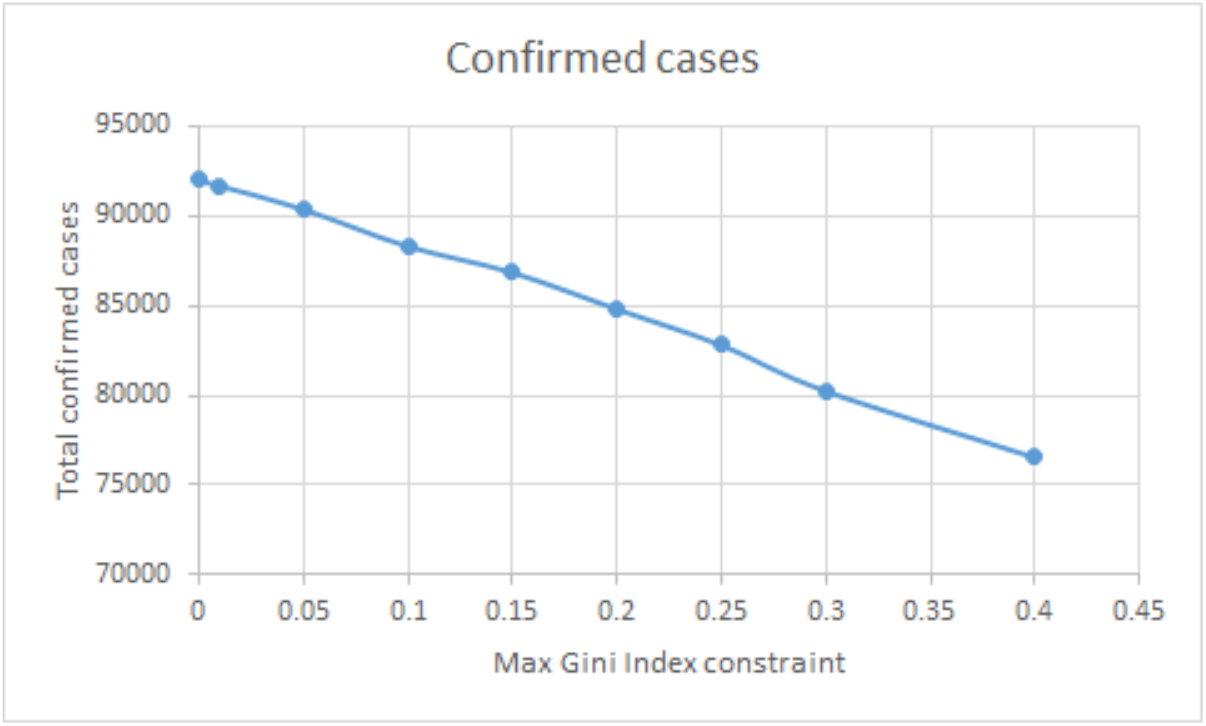
Confirmed cases under the myopic policy with the Gini index constraint

**Figure EC.2.**
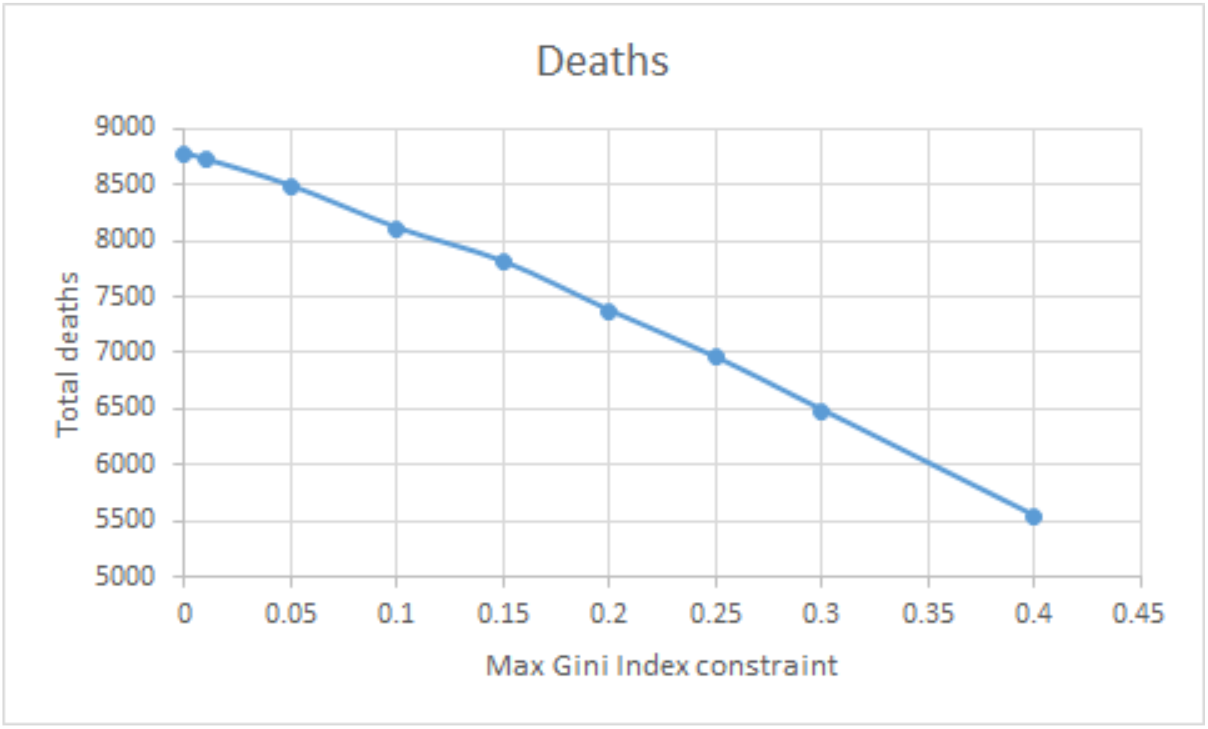
Deaths under the myopic policy with the Gini index constraint

### EC.2. Details of Parameter Estimation

In this appendix, we provide details of the parameter estimation method.

The input to our SAPHIRE model is simply the parameters and the observed initial conditions. Then the model can compute the daily number of each groups from previous days’ data with the equations defined in Section 3. The initial condition comes from the observation on Mar 17, 2020. We set the initial susceptible population to be the total population, compute the initial number of ascertained people from each group by the total infected number times the overall ratio of different age groups, and set the initial removed population to be 0. Note that the number of some disease group is not observable, for example, the unascertained infected people. However, we argue that as early as Mar 17, there is not too many people in this group. With our final set of parameters, if we multiply or divide the initial number in these unobserved compartments by 2, the change in our estimated total number of infected cases would be around 1.5%. To further deal with this, we set the first ten days in our model to be the burn-in period, and we will only take predictions of our model from Mar 27 to June 8.

**Figure EC.3.**
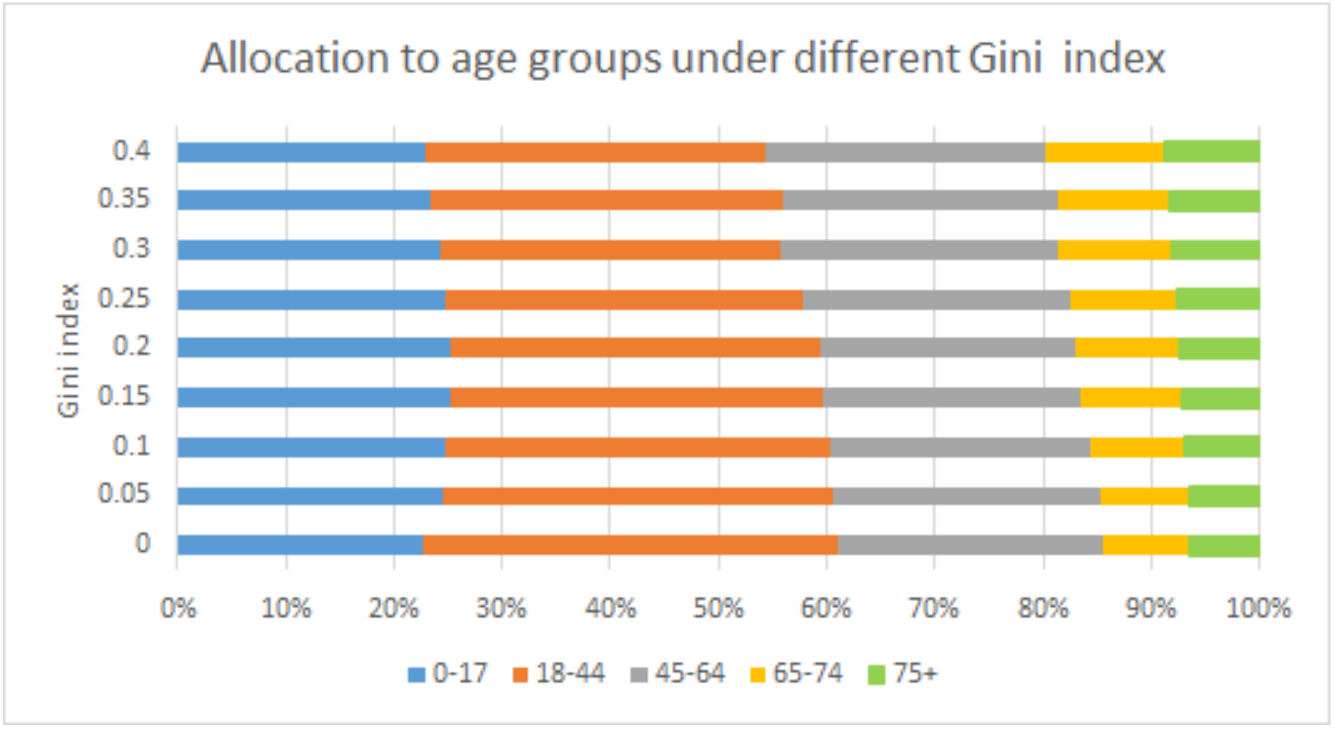
Fraction of total capacities allocated to each group

**Figure EC.4.**
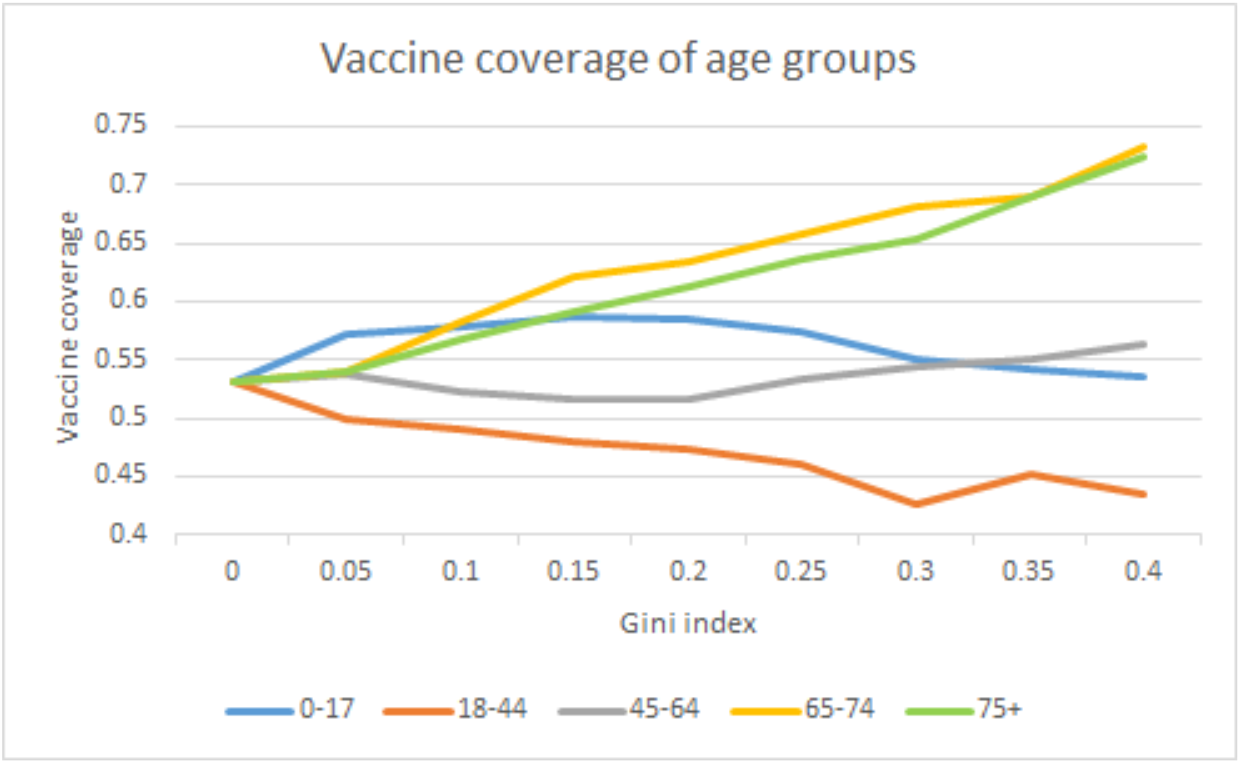
Vaccine coverage of each group

As we have mentioned before, to estimate the parameters, our loss function to be minimized is the squared error of the prediction of confirmed cases and deaths in each groups. Formally, let *x*(*t*) be the state vector at time t satisfying ODEs 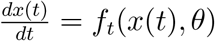 and *x^o^*(*t*) be the observable state subvector. Let *y*(*t*) be the observable data in time *t*. The loss function is written in (EC.3). This is a common measure used in the literature, see, for example, Canto et al. (2017). Due to the highly non-convex nature of the ODEs that define the epidemic dynamics, the loss function possess a great number of local optimal solutions, many of which have similar level of mean squared error in the prediction. These local optimal solutions can be obtained from applying optimization algorithm to the loss function from different starting points.

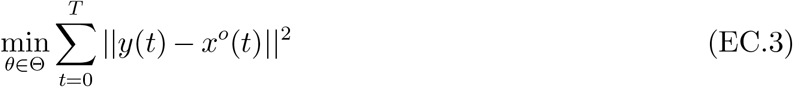

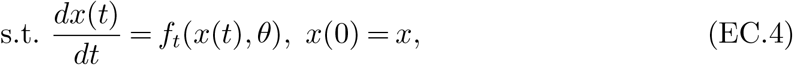

A critical part of our estimation is to select the local optimal solution that best characterizes the nature of transmission of the virus. When selecting among the multiple estimation of contact matrix *τ* between age groups, and the ascertainment rate *r* of different age groups, we consider the following constraints on the estimation.

First, the contact between age groups should be balanced and bi-directional. In other words, when two person makes contact, it is possible to transfer virus from the first person to the second person, or the reverse. In terms of contact between age groups, the following equation must hold for all elements in *τ*

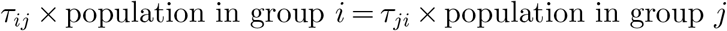

Second, there are some studies in finding the contact rate between people using socioeconomic data, for example, Fumanelli et al. (2012) uses the data of school, workplace and community to estimate the contact rate between age groups in the UK. Refer to Figure 2 of the paper for a heatmap of the estimated contact level between age groups. Though as a metropolitan area, people in New York City during the lock down period may not have exactly the same contact level as shown in this paper, but some insights should be similar. For example, the diagonal elements of the matrix *τ* should be the largest in a row, and the more off-diagonal an elements is in a row, the smaller it is. Formally, we incorporate the following constraint, with a small tolerance.

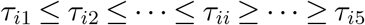

Third, as data have shown, the hospitalization rate and fatality rate of the older group is significantly higher than the younger group, we expect the ascertainment rate *r* for the groups follow the same fashion. In this sense, we incorporate the constraint that the ascertainment rate should be increasing in age. Formally,

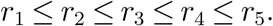

Based on the above criteria, we find the local minimum solution that satisfy the above constraints, that yields the smallest objective value. The results are shown in the main text.

